## Supplemental data for "Diagnostic Utility of Genome-wide DNA Methylation Analysis in Genetically Unsolved Developmental and Epileptic Encephalopathies and Refinement of a CHD2 Episignature"

**by LaFlamme et al.**

**Supplemental Phenotype data**

*This section has been removed based on MedRxiv guidelines. For access to this data, please contact the corresponding author.*

### Supplemental Materials and Methods

#### Cohort Subjects and Controls

Unaffected, presumably healthy controls without DEE include 111 healthy controls obtained through the Parkinson's Progression Markers Initiative (PPMI)[8], institutionally available data for 337 community control individuals without cancer from the St. Jude Life (SJLIFE) study[9], and 30 unaffected parents of participants in our study cohort (Table S1). Analytical controls include six individuals with known rare DMRs for validation of the computational outlier approach. These include two probands with Baratela-Scott syndrome (BSS) harboring *XYLT1* heterozygous hypermethylation on one allele and pathogenic variants on the other allele, one heterozygous *XYLT1* hypermethylation carrier parent of child with BSS, and three individuals with Fragile X syndrome harboring *FMR1* hypermethylation (two males, one female) obtained from the Coriell Institute for Medical Research (Table S1). Further analytical controls include 24 individuals with DEE caused by pathogenic variants in genes with known episignatures to validate the episignature screening, including n=15 *CHD2*, n=1 *CHD2* VUS, n=1 *KDM5C*, n=1 *SETD1B*, n=1 *KMT2A*, n=1 *SMARCA2*, n=2 *SMC1A*, and n=2 CNVs (Table S4). Finally, we included 116 analytical controls comprised of individuals with DEE who had previously identified pathogenic variants in genes without known episignatures. These 116 individuals better age match our unsolved DEE cohort and were used as further controls for comparison of rare DMRs and episignatures.

#### Reproducibility of DNA Methylation Array Replicates for Outlier Analysis

For individuals with replicate array data (n=29 individuals, n=61 total samples), we checked for the reproducibility of autosomal DMRs across different batches for the same individual. Of the 29 individuals, 17 of the individuals had perfect concordance across replicates for the DMRs that were called by the algorithm; six individuals had DMRs called for one replicate that could be confirmed through manual inspection. The remaining 6 individuals had at least 1 DMR that was not reproducible across replicates. Of 86 total DMRs for the 61 replicate samples, 18 did not reproduce across the replicates.

**Targeted Enzymatic Methyl-Sequencing Library Preparation, Target Enrichment, and Sequencing**

This approach uses two sets of enzymatic reactions to convert unmodified cytosines to uracil, allowing for the detection of methylated versus unmethylated cytosines without the need for bisulfite conversion[10].

Approximately 200-300ng of peripheral blood-derived genomic DNA input for 9 samples (three positive controls (*XYLT1* and *FMR1* hypermethylation) and three DEE probands with DMRs-of-interest) were diluted to 52μL with 0.1X TE pH 8.0 in a 96 microTUBE-50 AFA fiber plate (PN 520168) and fragmented on the Covaris LE 220 with SonoLab 7.3 software (Covaris Inc.). A 120-second program with peak incident power (W) of 453.0, duty factor (%) of 15.2, and cycles per burst (cpb) of 1000 resulted in an average fragment size of 285 bp. Then, 50μL of each sample was transferred to a fresh plate to begin library construction. End repair, A-tailing, and the ligation of the 0.4μM EM-Seq adaptor (A5mCA5mCT5mCTTT5mC5mC%mCTA5mCA5mCGA5mCG5mCT5mCTT5mC5mCGAT5mC*T and [Phos]GAT5mCGGAAGAG5mCA5mCA5mCGT5mCTGAA5mCT5mC5mCAGT5mCA) were performed with NEBNext DNA Ultra II reagents according to the manufacturer's instructions (PN 101977). Products were then purified by 1.18x bead clean up, using Twist Total Purification Beads (PN 101979) and eluted in 30μL water. About 28μL of each eluate was used as input for the 50μL oxidation reaction by TET2, proteinase K stop digestion, 1.8x bead clean-up, formamide denaturation, APOBEC deamination, and 1.0x bead purification described previously[10]. Then, 20μL of each deaminated library was used as input in an amplification reaction with 25μL of NEBNext Q5U Master Mix (#M0597) and 1μM (5μL) of primer from NEBs 96 Unique Dual Index

Primer Pairs Plate (#E7166A) as follows: 30 sec at 98°C; cycling 10 times, 10 sec at 98°C, 30 sec at 62°C, and 60 sec at 65°C; with a final extension for 5 minutes and hold at 4°C.

A 0.9x bead purification was performed on each amplified library, and 20µL of each eluate was used as input for target enrichment with the Twist Biosciences Fixed Human Methylome Panel. The 8-plex hybridization captures were created with 200ng of each sample combined to create a 1.6ug pool. Then, 4µL of Twist's Fixed Methylome Panel (PN 105521), 8µL of Universal Blockers, 5µL of Blocker Solution (#100767), and 2µL of Methylation Enhancer (#103558) were added to each pool and then dried down using an Eppendorf Vacufuge Plus with V-AQ setting @ 45°C for approximately 1 hour to create a pre-hybridization solution. Twist's Fast Hybridization mix (PN 104182) was heated at 65°C for at least 10 minutes until all precipitates dissolved, and 20µL was added directly to each of the dried pools without allowing the reagent to cool. Then, 30µL of Hybridization Enhancer was added to the top of each pool, and these pools were transferred to a thermal cycler set to a 95°C hold. Once the lid was sealed, a 95°C hold for 5 minutes was initiated, and samples were subsequently held overnight at 60°C for a minimum of 16 hours. Then, 100µL of Streptavidin Binding Beads (#100984) were washed with 200µL RT Fast Binding Buffer, placed on a magnet for 1 minute, supernatant removed, for a total of 3 washes, a final 200µL FBB was added, and the beads were resuspended by vortexing. Each hybridization pool (8-plex) was added directly from the 60°C thermocycler to a tube of washed, resuspended Streptavidin Binding Beads and mixed at RT on a rotisserie axel for 30 minutes. Pools were then pulse spun and placed on a magnet for 1 minute. The supernatants were removed and discarded. Tubes were removed from their magnets, and 200µL of preheated (62°C) Fast Wash Buffer 1 was added to each pool and pipette mixed. Pools were then incubated at 62°C for 5 minutes, pelleted on magnets for 1 minute, and the supernatant removed and discarded. Then, 200µL FWB1 addition and incubation were repeated, tubes were spun down, and bead-sample mixtures were transferred to fresh 1.5mL tubes before placing them on magnets. The remainder of the washes and final elution of enriched 8-plex libraries were performed according to the manufacturer's instructions (Twist Biosciences). Then, 30µL of each final enriched library was transferred to a clean tube for Illumina short-read sequencing.

All inputs and libraries were evaluated and quantified using D1000HS tape for TapeStation (Agilent) and Qubit™ 1X dsDNA HS (Invitrogen) assay kits. The average final library size was 370 bp. Sequencing was performed on the NovaSeq 6000 platform in paired-end mode with 150 bp per read. Paired-end reads (151 bp) for EM-Seq were collected from NovaSeq 6000 runsand then analyzed using bcl2fastq. The hg38_noAltHla_UCSC.fa reference file supplied through FTP by Twist Biosciences was used to create a reference genome in FASTA format. Samtools[11] and picard were used to create an index and ref sequence dictionary. The raw FASTQ data underwent quality control, processing and downstream analysis using the nf-core/methyseq bioinformatics pipeline. To extract the methylation calls using MethylDackel[12], we invoked the '—aligner bwameth' flag in the nf-core/methyseq pipeline. EM-Seq samples were trimmed using Trim Galore! (Krueger, Felix, 2015, v. 0.6.6) and cutadapt, and subsequently aligned using bwameth (default parameters), sambamba, and samtools. Duplicate reads were marked (picard) with an optical pixel distance of 2500 for a patterned S4 flow cell (Illumina). The covered_targets_Twist_Methylome_hg38_annotated_collapsed.bed file with TARGET and BAITS was converted to an interval list, and performance metrics were generated (fold-80 base penalty, HS library size, % duplicates, % off bait). MethylDackel was used to generate a methylation bias plot, likely variant sites were excluded (--maxVariantFrac 0.25), minimum depth of 10X (--minDepth 10) was set, and CpG calls were generated. Additional filters were added to generate global methylation cytosine reports, including CHH and CHG sites. Mapping efficiency, mapped reads (samtools), and the global average CpG % methylation and non-CpG conversion ratio (%) for each sample were generated from the cytosine report (methyldackel).

**Exome and Genome Sequencing**

Variants for individual samples were called from ES and GS following the GATK best practices workflow (including VQSR)[13]. Minimum genotype quality (≥20), minimum coverage depth (≥7), and minimum variant allele frequency (≥20%) thresholds were applied with Bcftools[11]. Variants were annotated with population frequency data (gnomAD[14], ExAC[15], ESP6500[16], and 1000 Genomes[17]), in silico scores (dbNSFP[18]), and potential clinical relevance (ClinVar[19] and InterVar[20]) via ANNOVAR[19] and InterVar[20]) via ANNOVAR[21]. Additional in silico scores (LOFTEE[22], CADD[23], and SpliceAI[22]) were applied using VEP[23].

**RNA-Sequencing and Gene Expression Analysis**

RNA was extracted from a flash-frozen cell pellet obtained from cultured fibroblasts using the Quick-RNA Miniprep Kit (Zymo Research). RNA was quantified using the Quant-iT RiboGreen RNA assay (ThermoFisher), and quality was checked by the 2100 Bioanalyzer RNA 6000 Nano assay (Agilent) or 4200 TapeStation High Sensitivity RNA ScreenTape assay (Agilent) before library generation. Libraries were prepared from total RNA with the TruSeq Stranded mRNA Library Prep Kit according to the manufacturer's instructions (Illumina PN 20020595). Libraries were analyzed for insert size distribution using the 2100 BioAnalyzer High Sensitivity kit (Agilent), 4200 TapeStation D1000 ScreenTape assay (Agilent), or 5300 Fragment Analyzer NGS fragment kit (Agilent). Libraries were quantified using the Quant-iT PicoGreen ds DNA assay (ThermoFisher) or by low-pass sequencing with a MiSeq nano kit (Illumina). Paired-end 100-cycle sequencing was performed on a NovaSeq 6000 (Illumina). The raw FASTQ data underwent processing and analysis utilizing the nf-core/rnaseq bioinformatics pipeline[24].

**Whole-Genome Bisulfite Sequencing**

The EZ-96 DNA Methylation-Gold MagPrep (Zymo Research) was used for bisulfite conversion. Libraries were prepared from converted DNA using the xGen Methylation-Sequencing DNA Library Preparation Kit (Integrated DNA Technologies). Libraries were analyzed for insert size distribution using the 2100 BioAnalyzer High Sensitivity kit (Agilent), 4200 TapeStation D1000 ScreenTape assay (Agilent), or 5300 Fragment Analyzer NGS fragment kit (Agilent). Libraries were quantified using the Quant-iT PicoGreen ds DNA assay (ThermoFisher) or by low-pass sequencing with a MiSeq nano kit (Illumina). Paired-end 150-cycle sequencing was performed on a NovaSeq 6000 (Illumina).

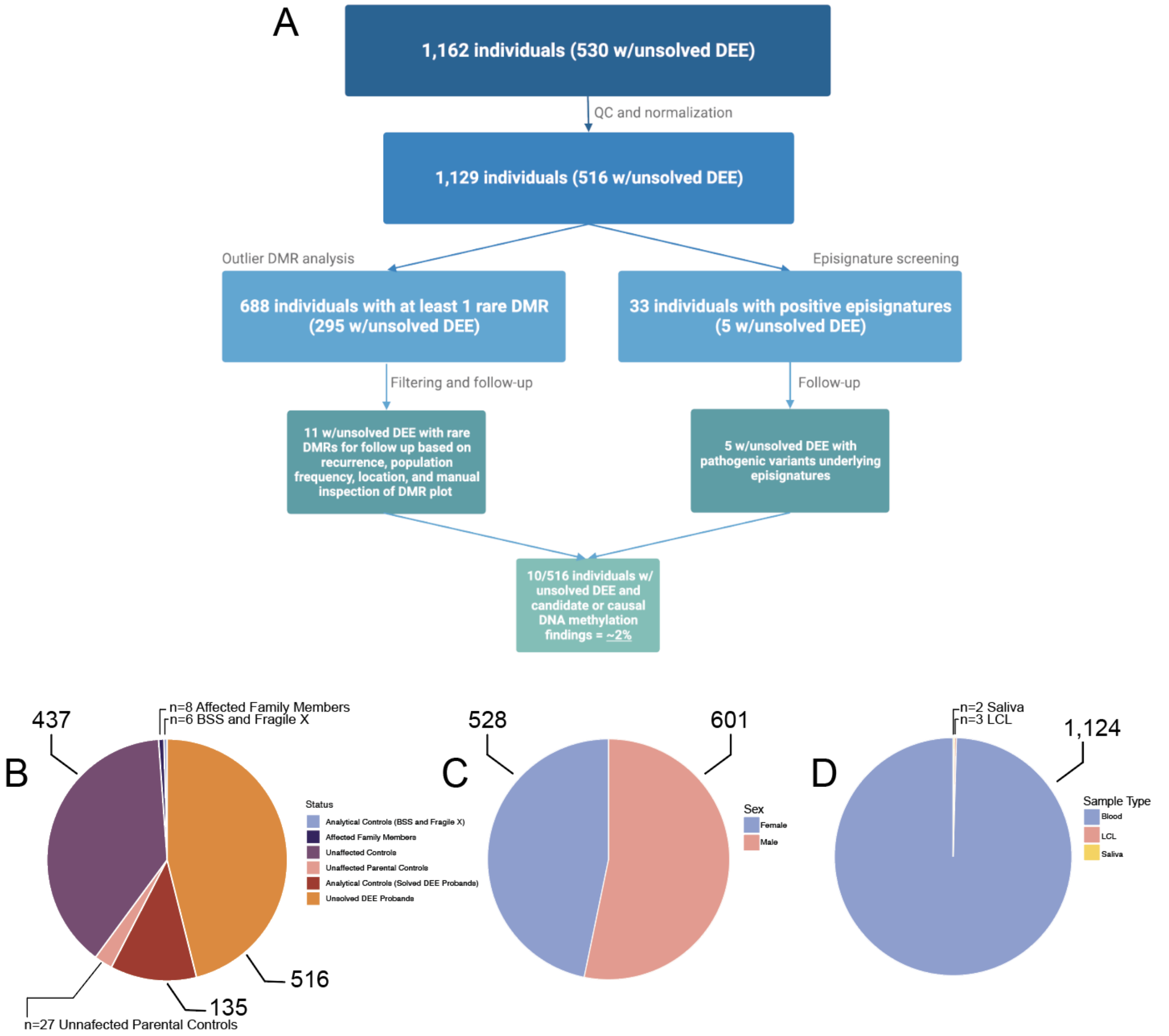


**Figure S1**. Flowchart of the methylation analysis and features of the study cohort. **A**. Flowchart displaying filtering of samples after quality control (QC) and normalization, analysis pipeline for detecting rare DMRs and episignatures, and narrowing down DNA methylation candidates for this study. Collectively, we find causal or candidate etiologies in 10/516 individuals with unsolved DEE using DNA methylation analysis. **B.** Breakdown of the cohort after QC and normalization (n=1,129) containing individuals with unsolved DEE (n=516), unaffected controls (n=464), analytical controls (n=141), and affected family members (n=8). **C.** Number of males (n=601) versus females (n=528). **D.** Sample type as blood (n=1,124) versus other tissue types (n=5).

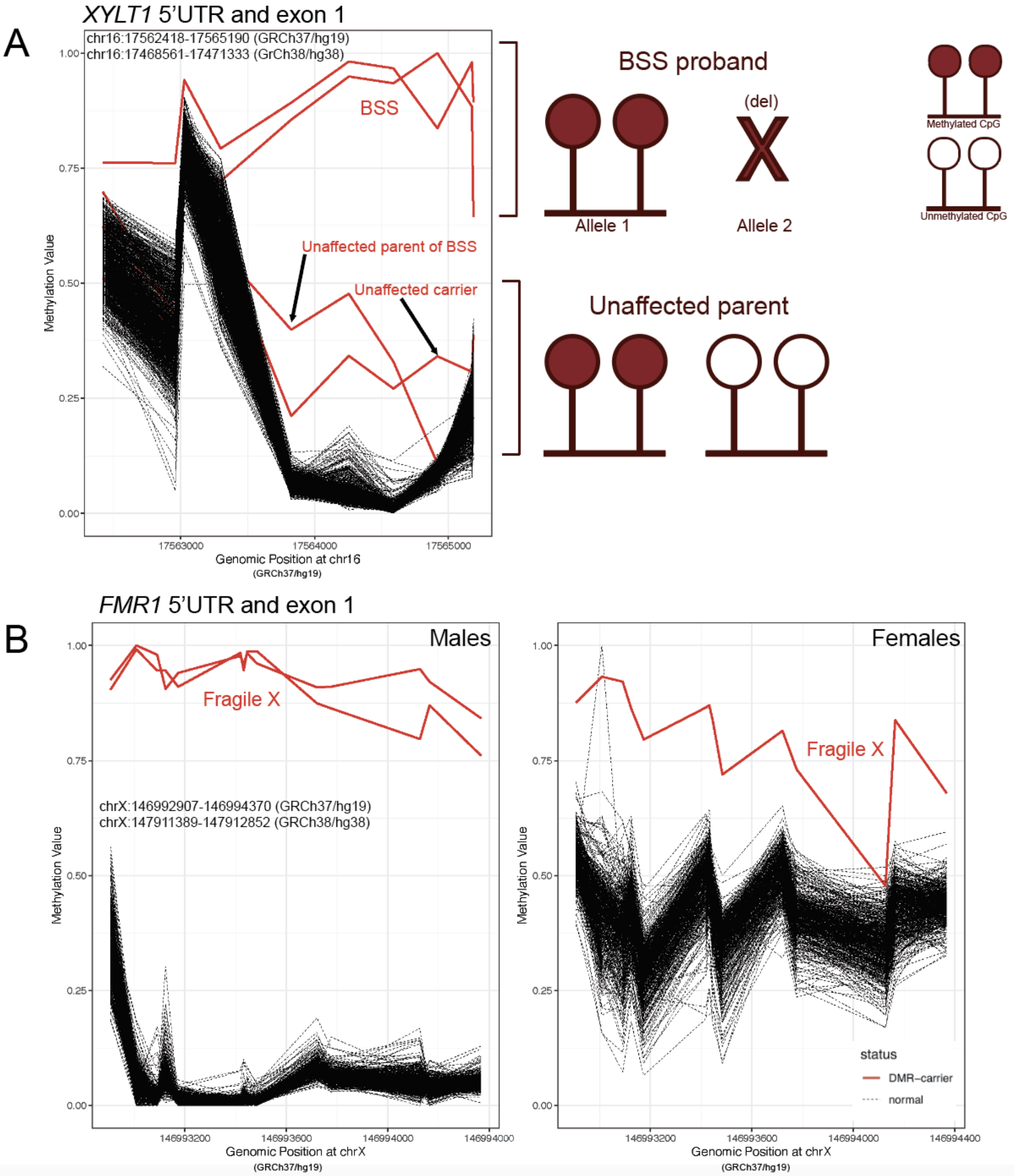


**Figure S2**. DMR plots from rare outlier analysis of known hypermethylation events. **A.** DMR plot of the *XYLT1* 5'UTR and exon 1 showing two individuals with Baratela-Scott syndrome (BSS) with hemizygous hypermethylation (red) around β=~1 compared to controls (black). Each inflection point corresponds to an individual CpG probe from the array. These individuals with BSS harbor hypermethylation of *XYLT1* on one allele and a deletion encompassing this region on the other, represented in the schematic to the right. One unaffected parent and another carrier (red) carry heterozygous hypermethylation around $\beta$=0.5. **B.** DMR plots of the *FMR1* 5'UTR and exon 1 with individuals split by males (left) and females (right) for targeted sex chromosome outlier DMR analysis. Fragile X males and females are clearly hypermethylated ($\beta$=~1) compared to their counterpart controls.

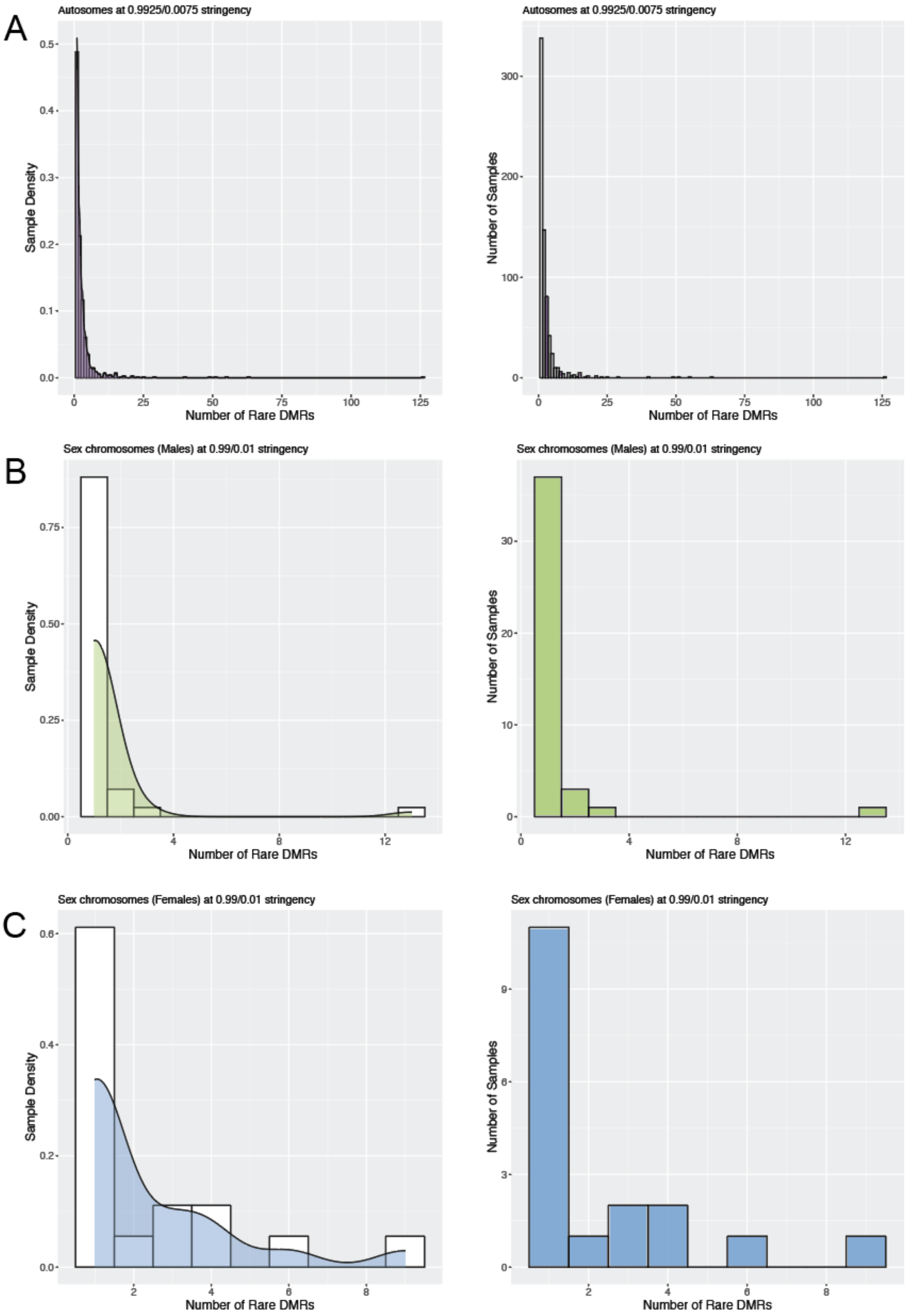
A
Autosomes at 0.9925/0.0075 stringency
Sample Density
Number of Rare DMRs
Autosomes at 0.9925/0.0075 stringency
Number of Samples
Number of Rare DMRs
B
Sex chromosomes (Males) at 0.99/0.01 stringency
Sample Density
Number of Rare DMRs
Sex chromosomes (Males) at 0.99/0.01 stringency
Number of Samples
Number of Rare DMRs
C
Sex chromosomes (Females) at 0.99/0.01 stringency
Sample Density
Number of Rare DMRs
Sex chromosomes (Females) at 0.99/0.01 stringency
Number of Samples
Number of Rare DMRs

**Figure S3.** Distribution of outlier DMRs across samples. Distribution plots showing sample density (left) and sample counts (right). **A.** Autosomes are in purple for the full cohort (n=1,156 array samples), **B.** chrX for male samples (n= 615 array samples) are in green, and **C.** chrX for female samples (n= 541 array samples) are in blue.

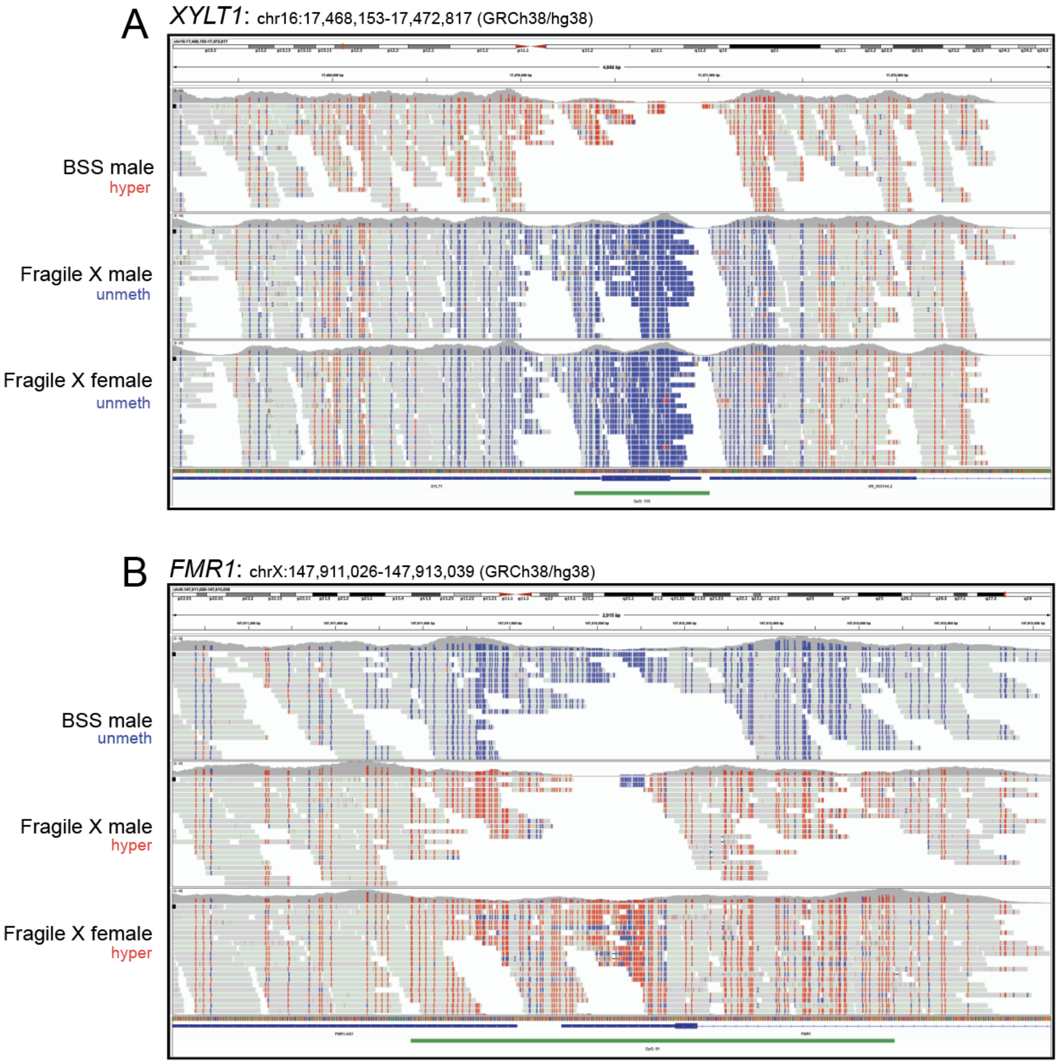


**Figure S4.** Validation of known outlier DMRs using targeted EM-seq. **A.** IGV view of targeted EM-seq reads at the *XYLT1* 5'UTR and exon 1 that are hypermethylated in an individual with BSS (upper) compared to methylation-negative controls (middle and lower). **B.** IGV screenshot of targeted EM-seq reads at the *FMR1* 5'UTR and exon 1 hypermethylated in Fragile X male and female individuals (middle and lower) compared to methylation-negative control (upper).

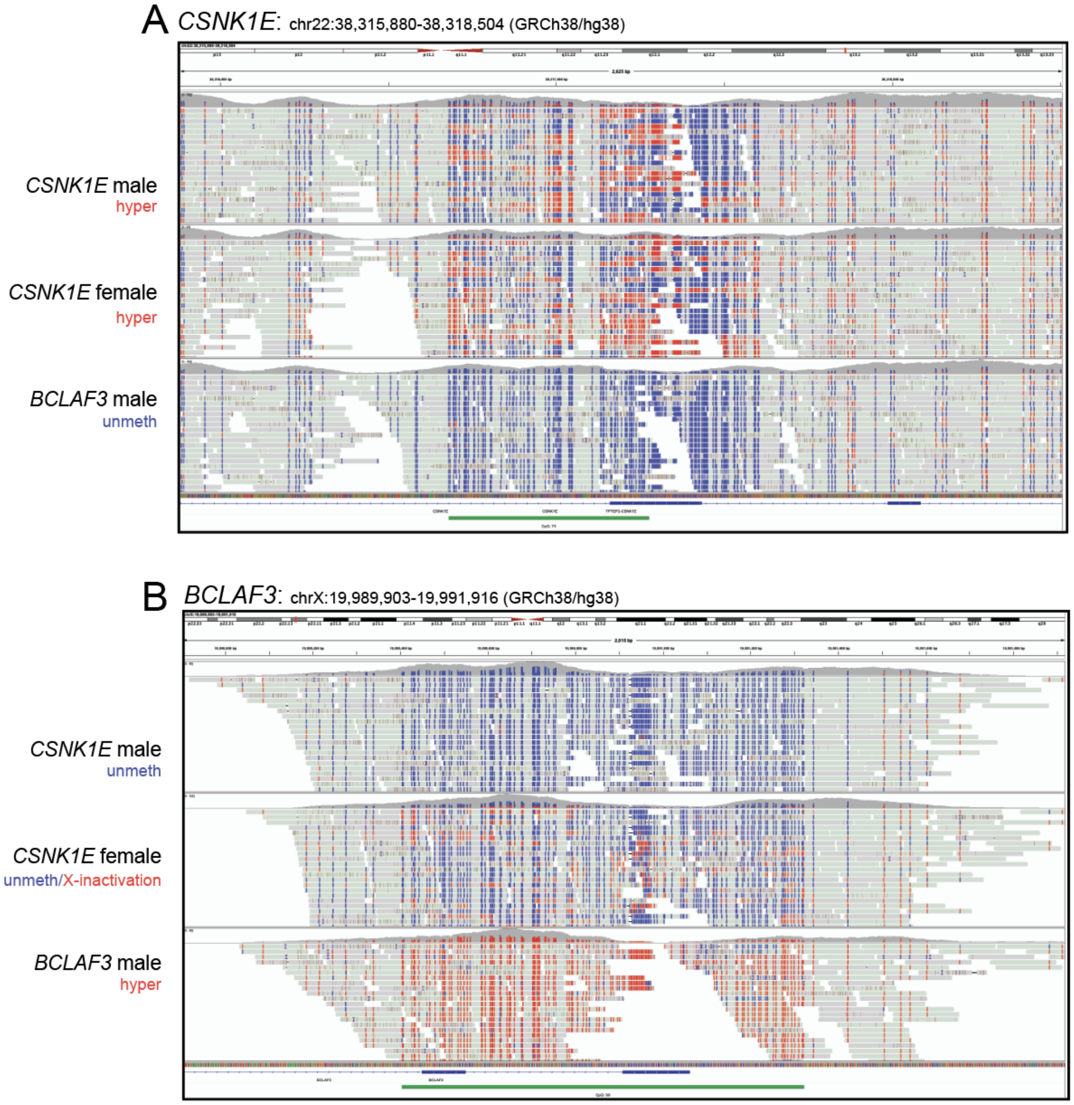


**Figure S5.** Validation of rare outlier DMRs called from methylation array in individuals with unsolved DEE using targeted EM-seq. **A.** IGV view of *CSNK1E* 5'UTR and intron 1 validating heterozygous hypermethylation in two individuals with unsolved DEE (upper and middle) compared to methylation-negative control (lower). **B.** IGV view of *BCLAF3* 5'UTR validating hemizygous hypermethylation called from methylation array in a male individual with unsolved DEE (lower) compared to methylation-negative controls (upper and middle).

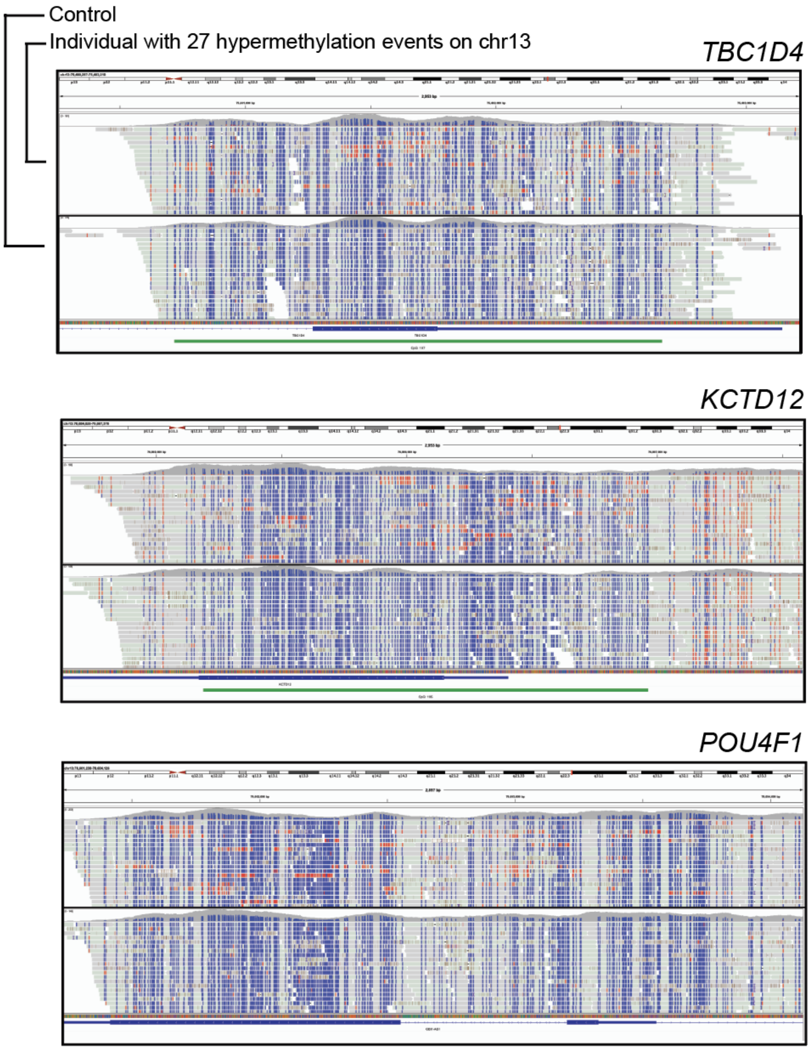


**Figure S6.** Validation of rare outlier DMRs on chr13 in a single individual using targeted EM-seq. Representative images depict IGV views for 3/27 DMRs on chr13 for an individual with unsolved DEE (upper panel of each box) compared to a control (lower panel of each box).

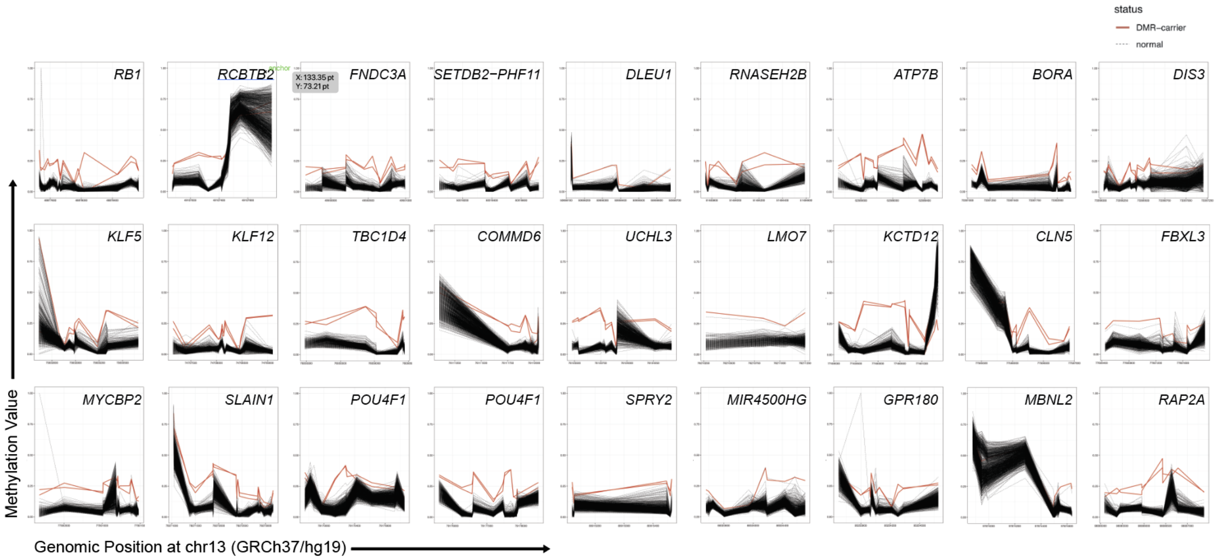


**Figure S7.** Individual with unsolved DEE displaying multiple hypermethylated DMRs across chr13. DMR plots depicting DNA methylation array data are shown for all 27 outlier DMRs called for an individual with unsolved DEE (red) compared to controls (black).

| | Family 1 | Family 2 | Family 3 |
|---|---|---|---|
| Methylation array (Blood) 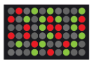 | inherited hypermethylation | *de novo* hypermethylation | inherited hypermethylation |
| Long-read sequencing (Blood) 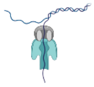 | 1,500-3,000bp CGG repeat expansion in the proband inherited from parent (~1,500bp) | NA | 1,300-2,100bp CGG repeat expansion in proband inherited from parent (~270-3,500) |
| RNA-seq (Fibroblasts) 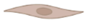 | NA | Decreased gene expression in proband | Decreased gene expression in proband and parent |

**Figure S8.** Table shows the data types analyzed from each family and a summary of any associated results.

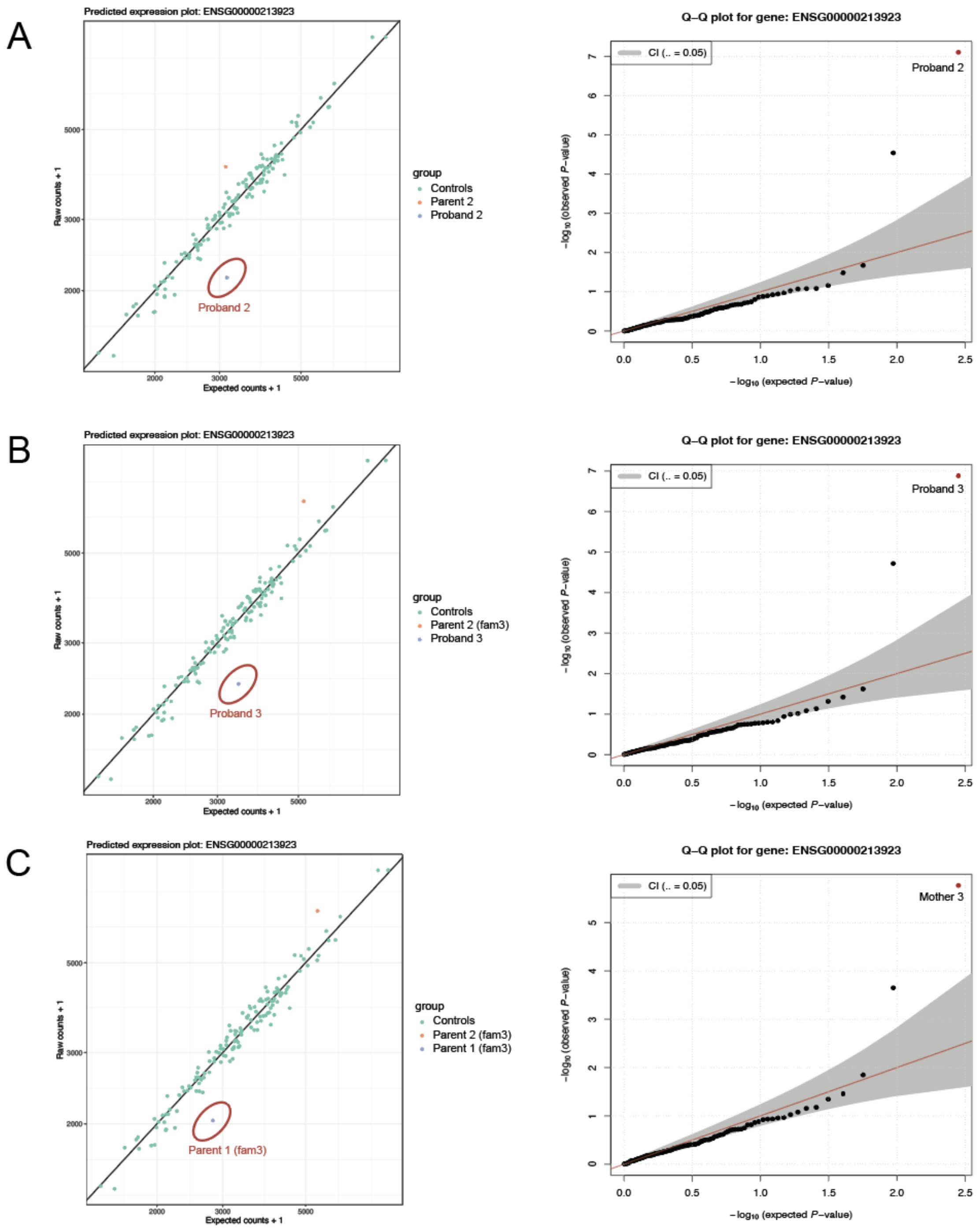


**Figure S9.** Dropout Analysis of RNA-seq data for *CSNK1E* from human-derived fibroblasts by family. OUTRIDER predicted expression plot is shown on the left and Q-Q plot on the right. **A**. Proband 2 "drops out" for *CSNK1E* expression (*padj*=0.00112276, zScore=-5.63, $\log_2$FC=-0.55) compared to Parent 2 and control fibroblast cohort of 139 publicly available samples[25]. **B**. Proband

3 “drops out” for *CSNK1E* expression compared to Parent 2 from Family 3 and the control fibroblast cohort (*padj*=0.00228623, zScore=-5.54, $\log_2$FC=-0.53). **C**. Parent 1 from Family 3 “drops out” for *CSNK1E* expression compared to Parent 2 from Family 3 3 and the control fibroblast cohort (*padj*=0.021391407, zScore=-5.02, $\log_2$FC=-0.47).

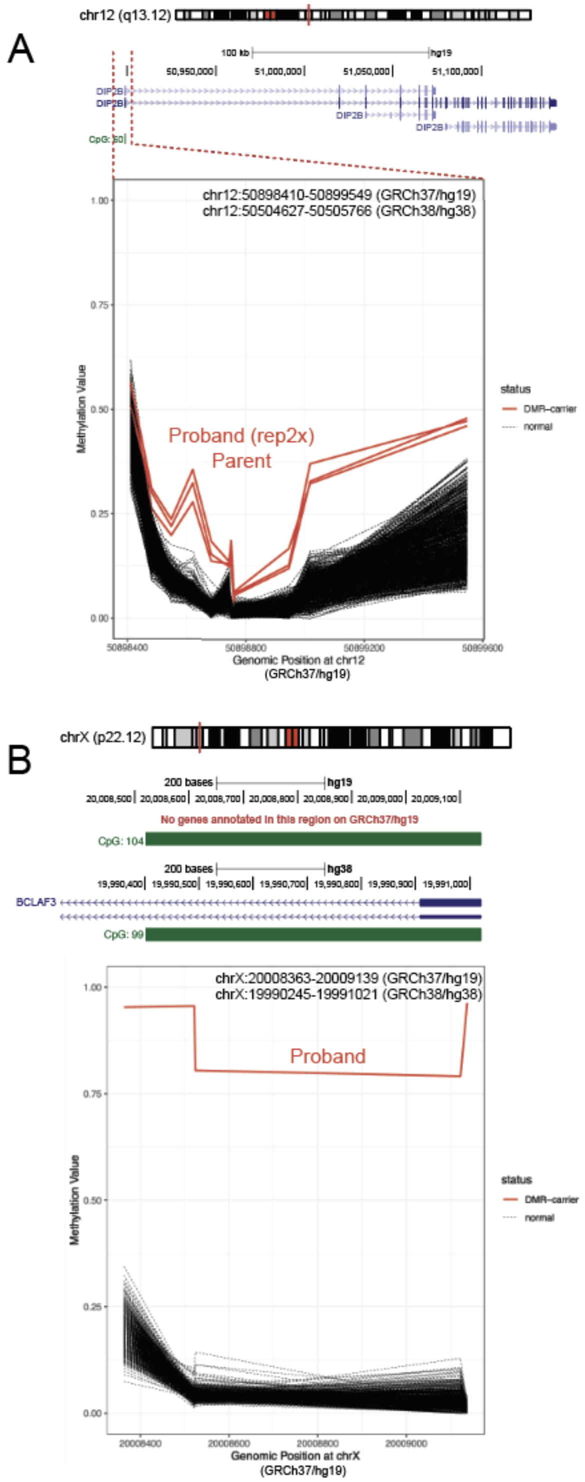


**Figure S10.** Outlier hypermethylation of regions underlying known and novel repeat expansions.

**A**. DMR plot for outlier hypermethylated DMR at the *DIP2B* promoter and exon 1 in a proband with unsolved DEE (repeated twice on the methylation array) and his parent (both in red). **B**. DMR plot of targeted X chromosome analysis in males depicting outlier hypermethylated DMR at an intergenic region (by GRCh37/hg19 annotation) and exon 1 of an uncharacterized gene *BCLAF3* (by GRCh38/hg38 annotation) in a male proband with unsolved DEE.

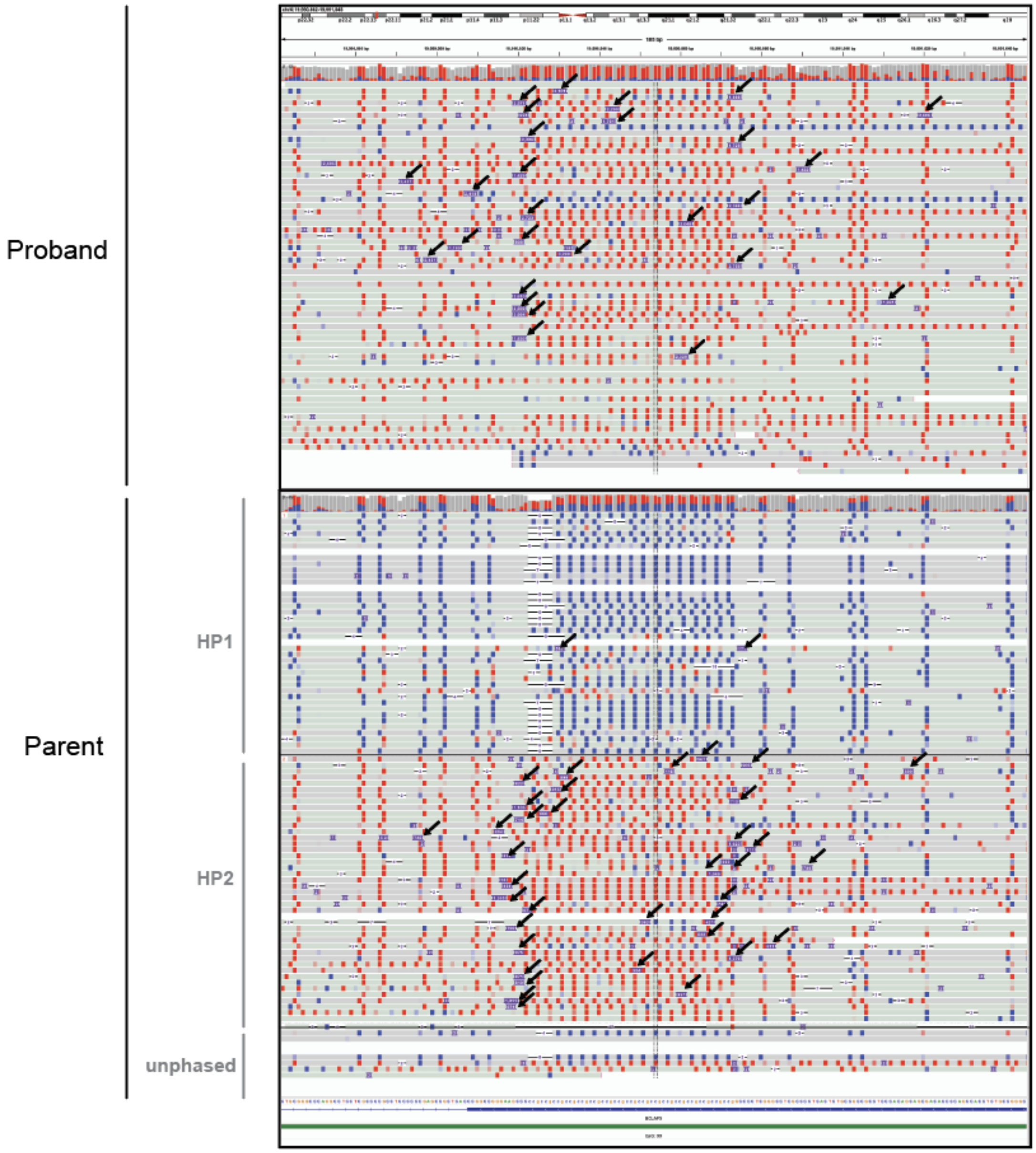


**Figure S11.** Long-read sequencing identifies a novel repeat expansion at the 5'UTR of *BCLAF3*. ONT long-read sequencing reads in IGV phased for 5mC for the proband validating *BCLAF3* hypermethylation (top panel). Long-read sequencing further indicates a novel ~2,500-3,000bp CG-rich repeat expansion (arrows). Haplotype resolved ONT long-read sequencing reads in IGV for the proband's parent (bottom panel) displaying smaller ~700-1,000bp CG-rich repeat expansion (arrows). Haplotype resolution also indicates that the parent's X-chromosome inactivation is skewed against the repeat allele, not a random 50/50 hypermethylation across both X chromosomes.

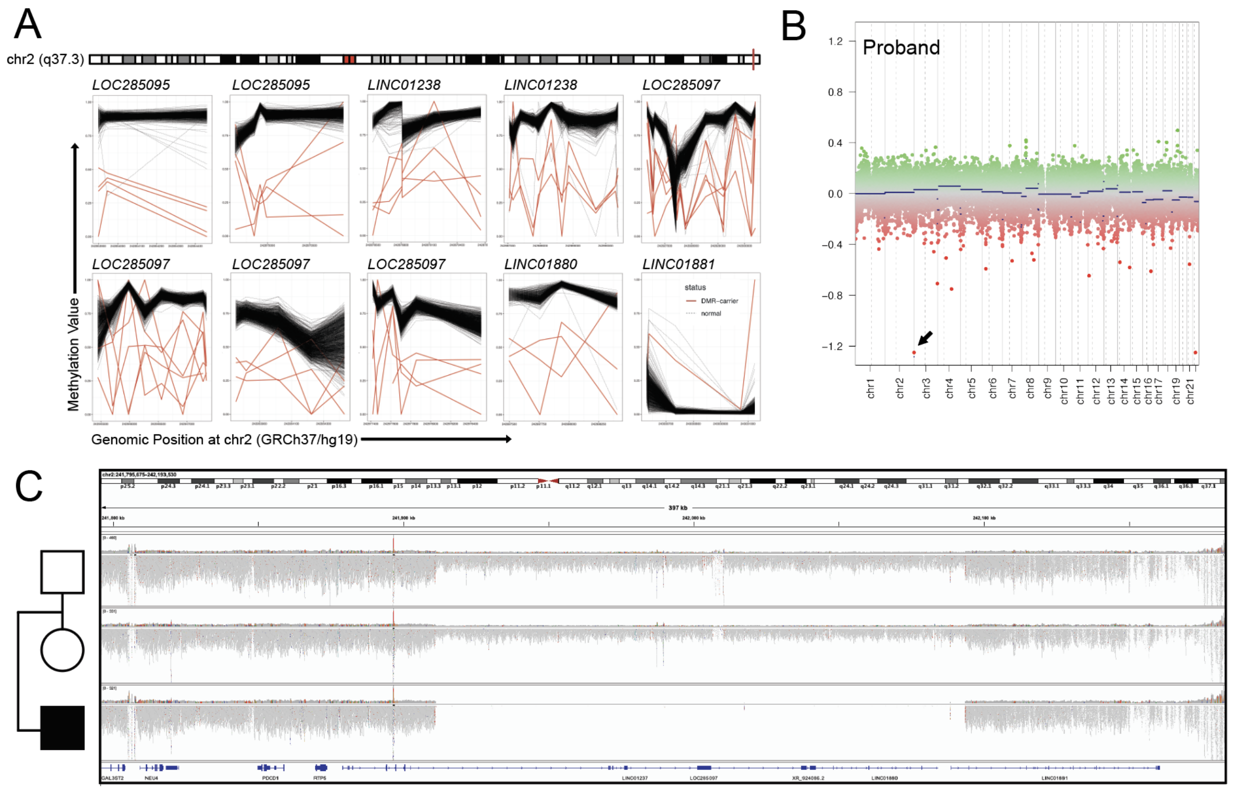


**Figure S12.** Outlier "DMRs" at chr2 represent copy number deletion. **A**. DMR plots showing "hypomethylated DMRs" across 2q37.3 for an individual with unsolved DEE. **B**. CNV calling from the array with "conumee" detects copy number deletion overlapping with chr2 DMR locus. **C**. GS of trio identifies inherited homozygous copy number deletion in proband.

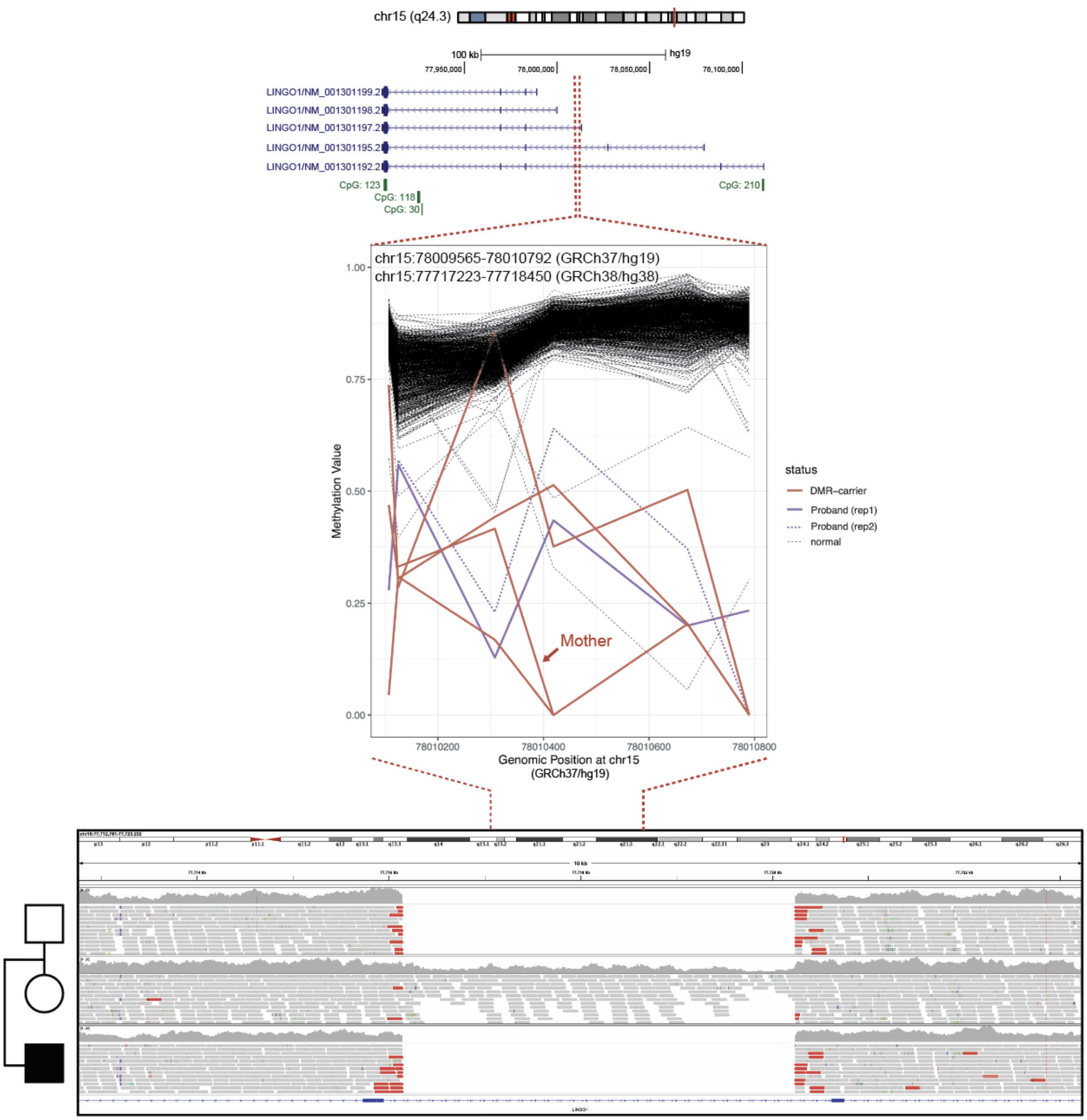


**Figure S13.** Outlier "DMR" at *LINGO1* intron represents copy number deletion. Annotation track showing the location of DMR in different *LINGO1* transcripts (upper). DMR plot showing hypomethylated DMR detected in several DMR carriers, including a proband with replicates (purple solid and dashed, middle). GS detects inherited copy number deletion in proband (lower).

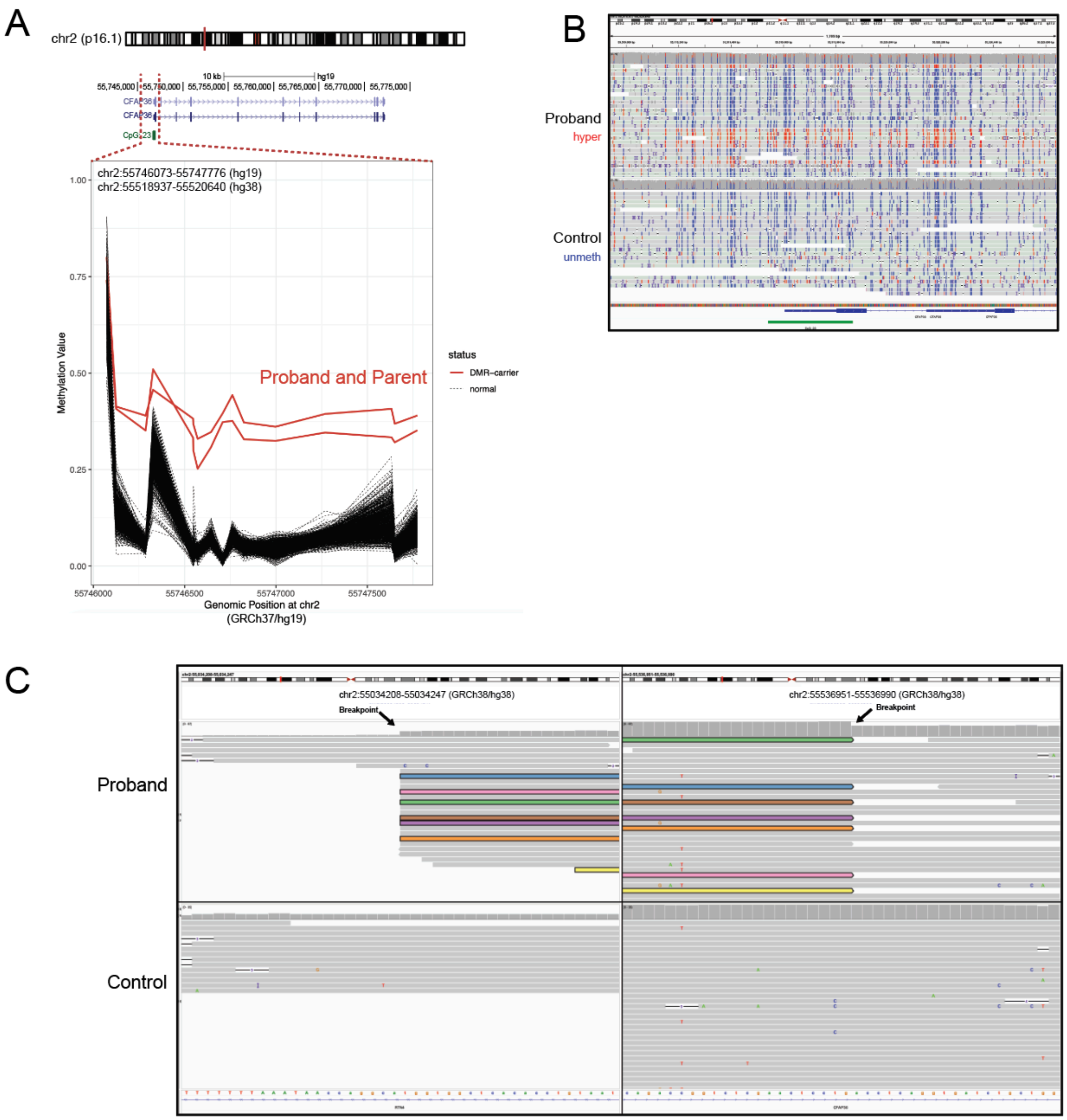


**Figure S14.** Outlier hypermethylated DMR at *CFAP36* due to a tandem duplication. **A**. DMR plot showing hypermethylation at CFAP36 in a proband with unsolved DEE and parent. **B**. IGV screenshot of targeted ONT long reads phased for 5mC at DMR region validating hypermethylation called from the array. **C**. IGV screenshot showing the location of the breakpoints of the tandem duplication and associated “dip” in coverage. Some but not all the reads showing the breakpoint are colored. The upstream breakpoint (left) was not included in the adaptive sampling target region and therefore has lower coverage compared to the target region.

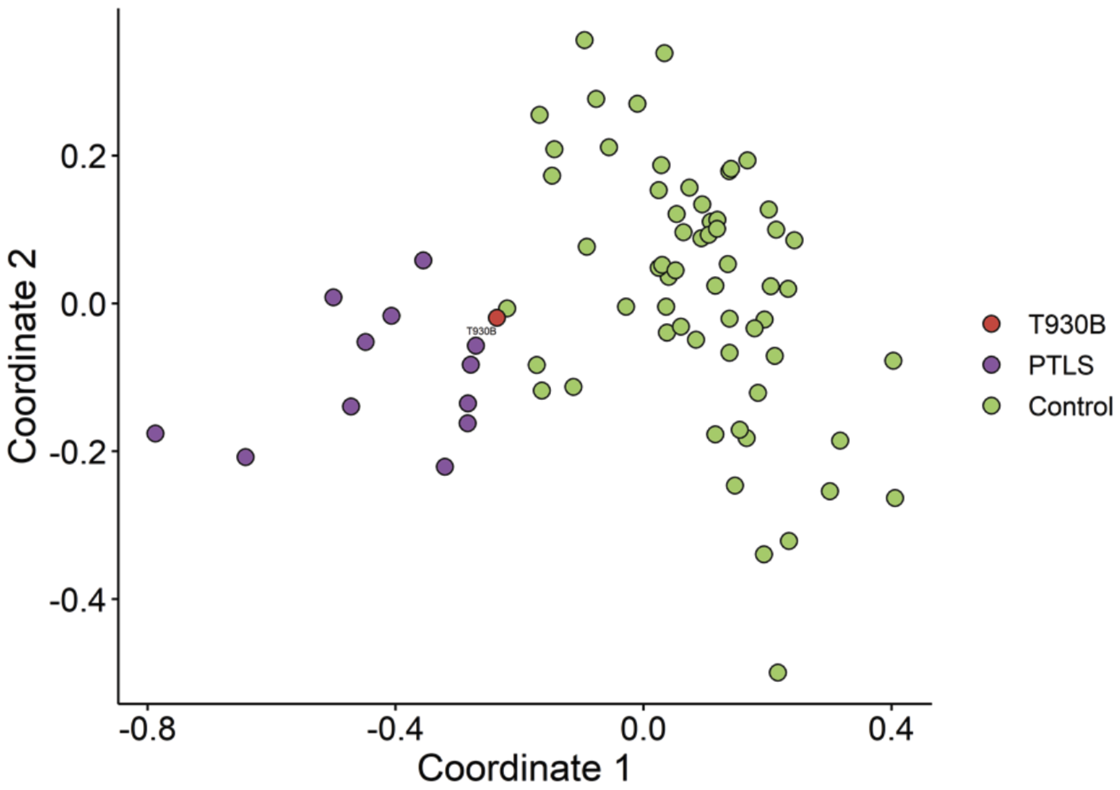


**Figure S15**. Inconclusive episignature results. An example of inconclusive results for Potocki-Lupski syndrome (PTLS) episignature. Individual T930B (red) was reported as inconclusive for PTLS due to a low MVP score of 0.053 and inconsistent MDS clustering between cases (purple) and unaffected controls (green). Inconclusive episignature results are reported with the caveat that further follow up or investigation may be warranted if there is a clinical phenotype consistent with the inconclusive episignature in question.

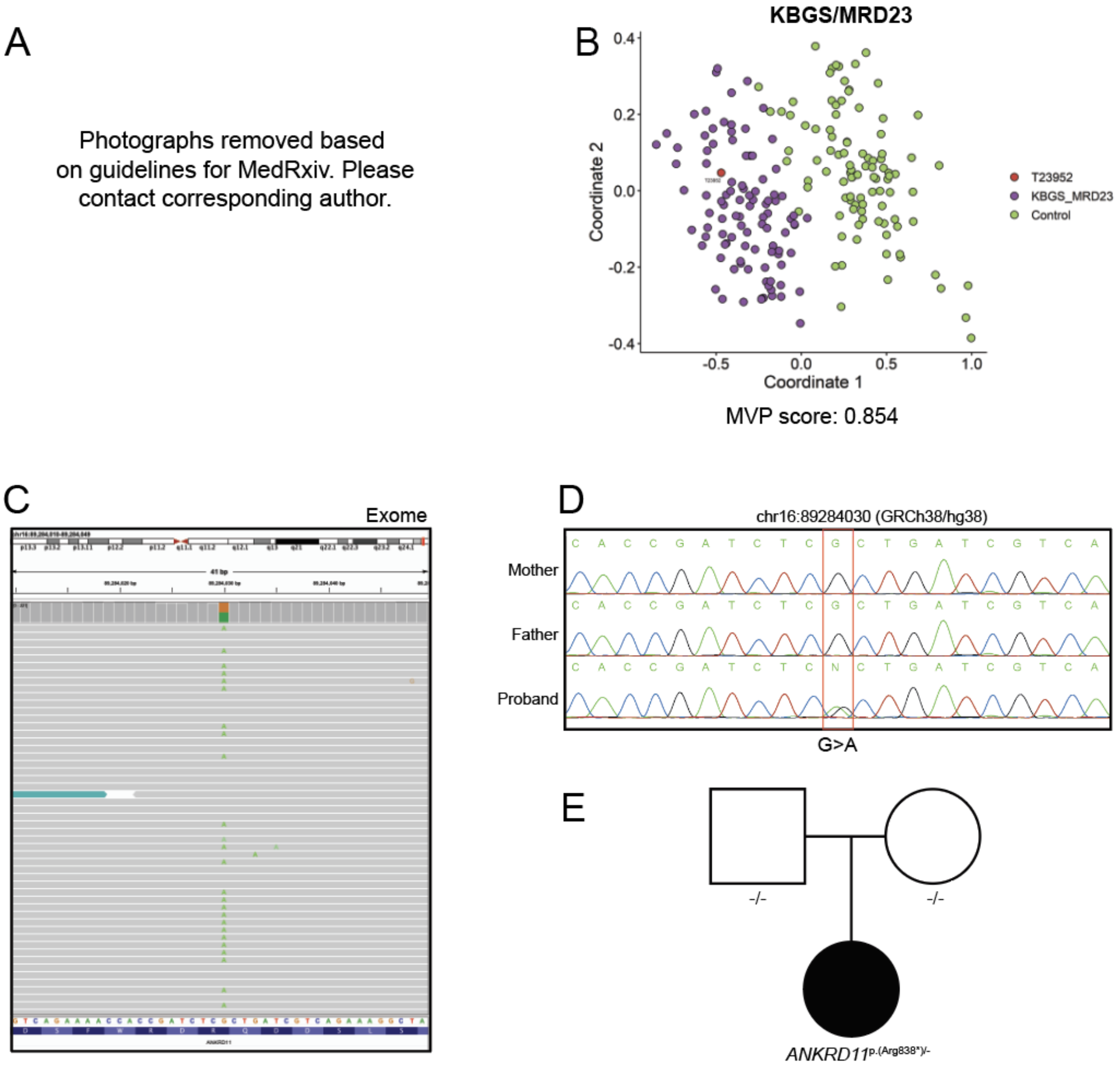


**Figure S16**. Diagnosis of *ANKRD11* enabled by episignature screening. **A.** Photos of affected individual displaying dysmorphic features, including triangular face, bulbous nose, thin upper lip, featureless philtrum, broad bushy eyebrows, large prominent ears, thin upper lip. **B.** Multidimensional scaling (MDS) plot showing how proband (red) clusters with KGBS_MRD23 episignature (purple). **C.** IGV screenshot of pathogenic variant in *ANKRD11* detected by exome sequencing. **D**. PCR validation and segregation of the variant. **E.** Pedigree indicating that the variant occurs *de novo*.

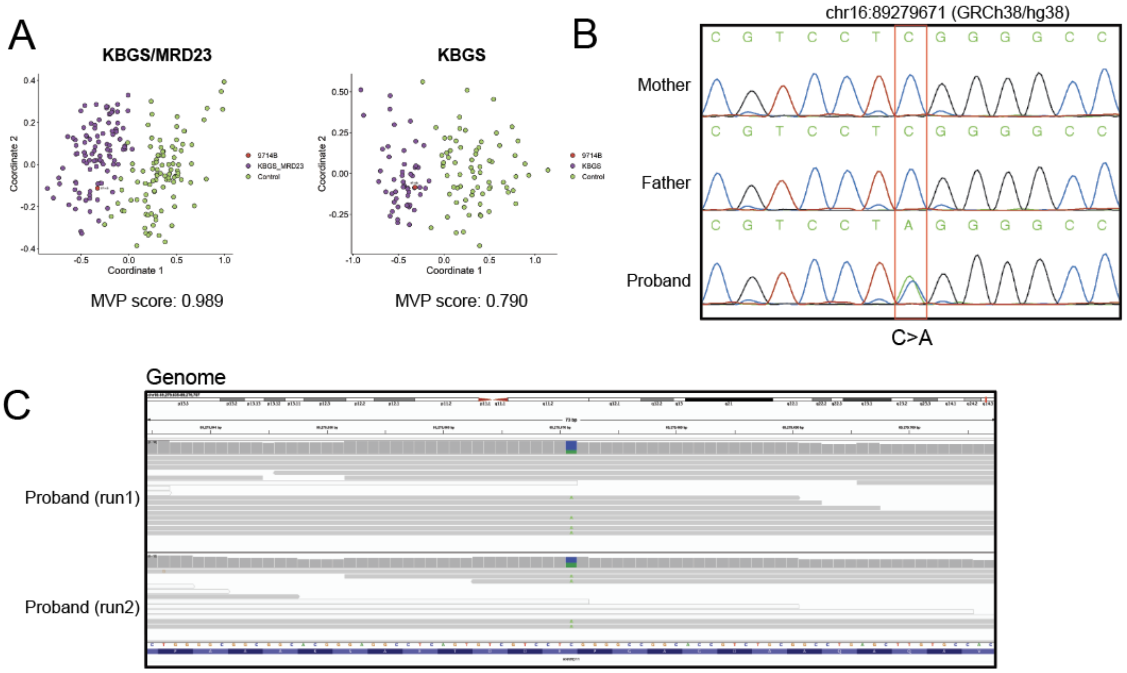


**Figure S17**. Diagnosis of *ANKRD11* enabled by episignature screening. **A**. MDS plot showing proband (red) clustering with KBGS_MRD23 affected controls (purple, left) and further clustering with the secondary KBGS signature (purple, right). **B**. Sanger sequencing validation and segregation of pathogenic variant recovered from **C**. Two low coverage GS runs showing the presence of the pathogenic variant. Since variant is *de novo* and not present in any of the tested affected family members, *ANRKRD11* variation is not the cause of the familial epilepsy but explains the more severe phenotype of the proband.

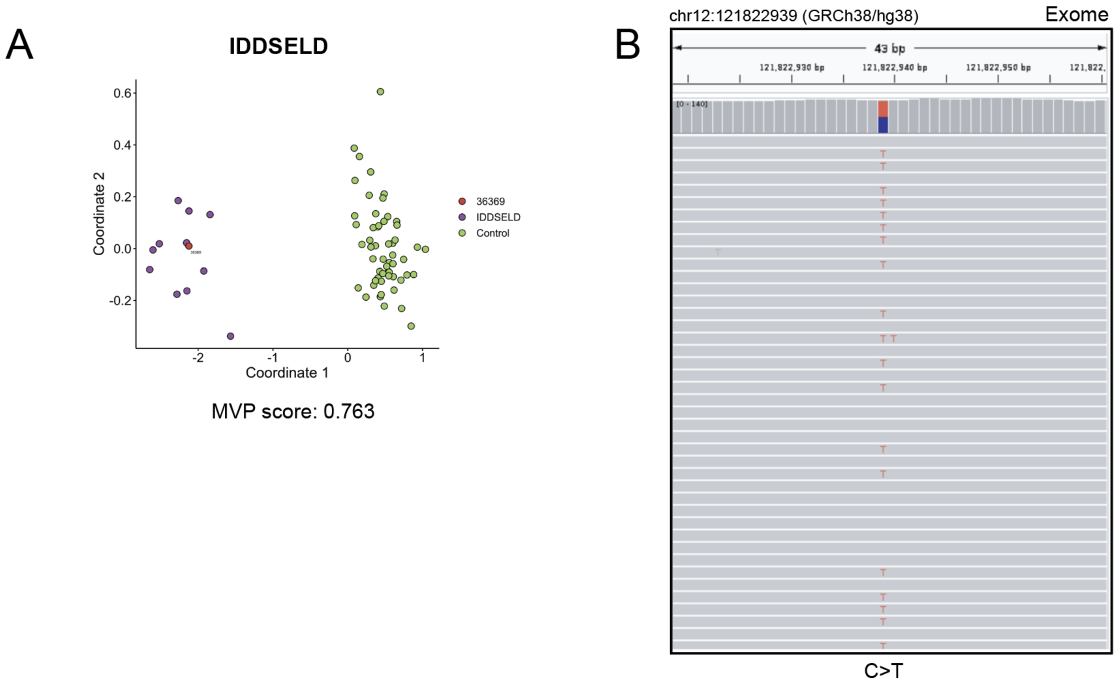


**Figure S18.** Diagnosis of *SETD1B* enabled by episignature screening. **A**. MDS plot showing clustering of the proband (red) with the IDDSELD episignature (purple). **B**. IGV screenshot of pathogenic variant identified by exome sequencing.

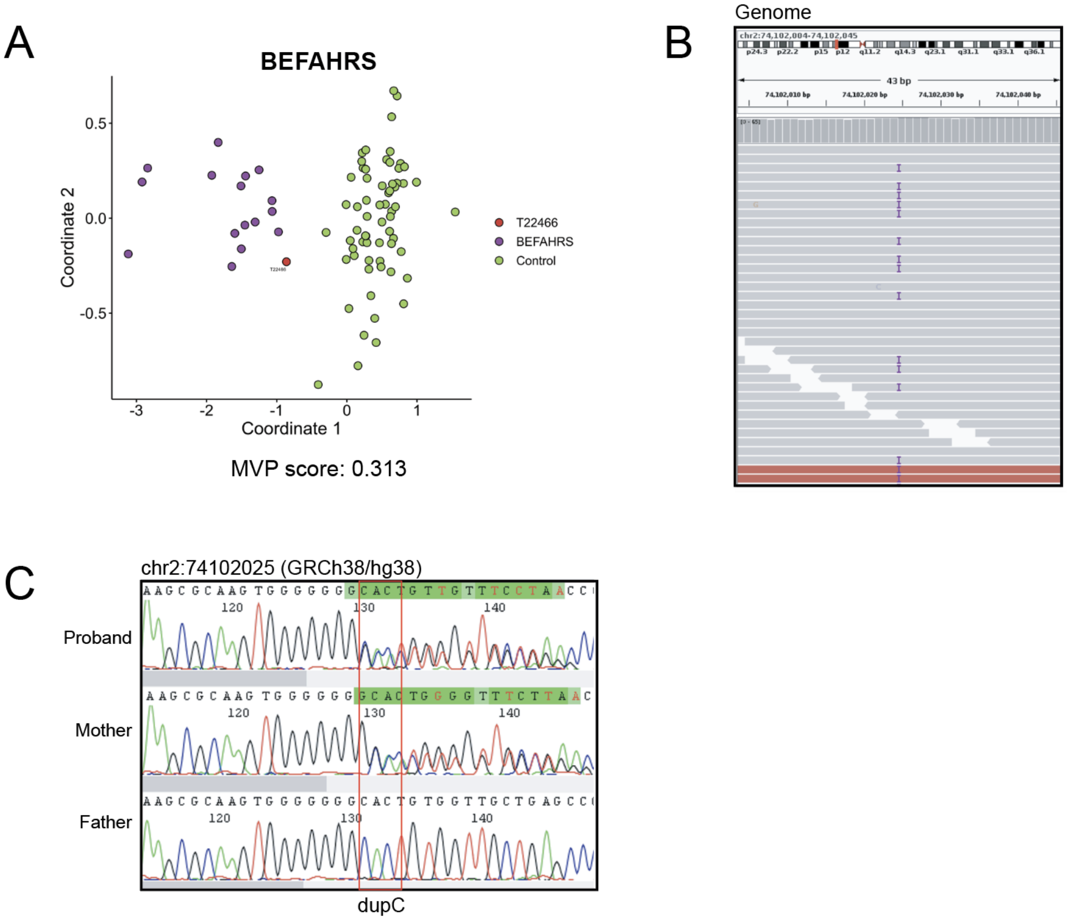


**Figure S19.** Diagnosis of *TET3* enabled by episignature screening. **A**. MDS plot showing clustering of the proband with the BEFAHRS episignature. **B**. IGV screenshot of pathogenic duplication identified through GS. **C**. Sanger sequencing validation and segregation of variant indicating that the *TET3* variant is inherited from the parent, who is mildly phenotypically affected (Supplemental Phenotype data).

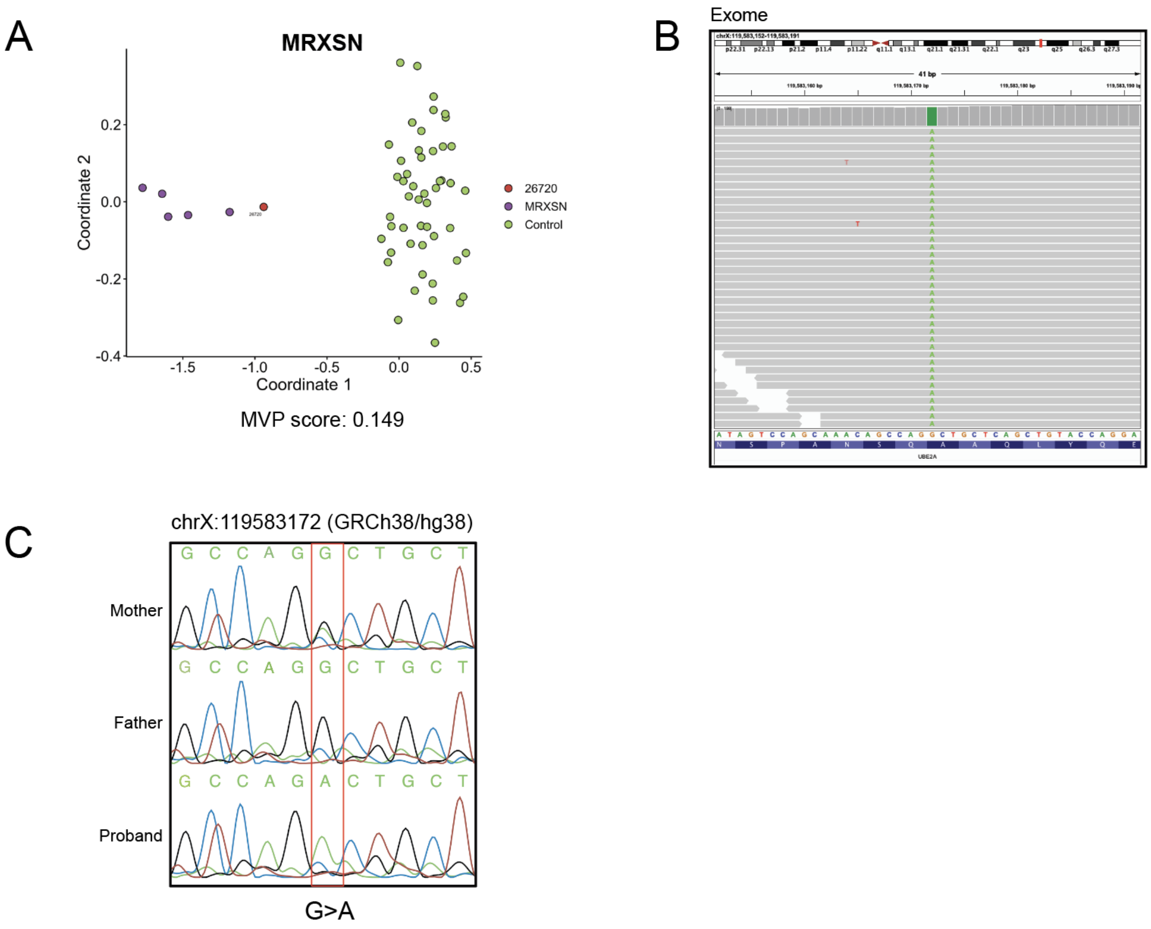


**Figure S20.** Diagnosis of *UBE2A* enabled by episignature screening. **A**. MDS plot showing clustering of the proband with the MRXSN episignature. **B**. IGV screenshot of hemizygous variant of uncertain significance identified by exome sequencing. **C**. Sanger sequencing validation and segregation of variant, determined to X-linked and inherited. Variant was determined to be pathogenic by clinical workup.

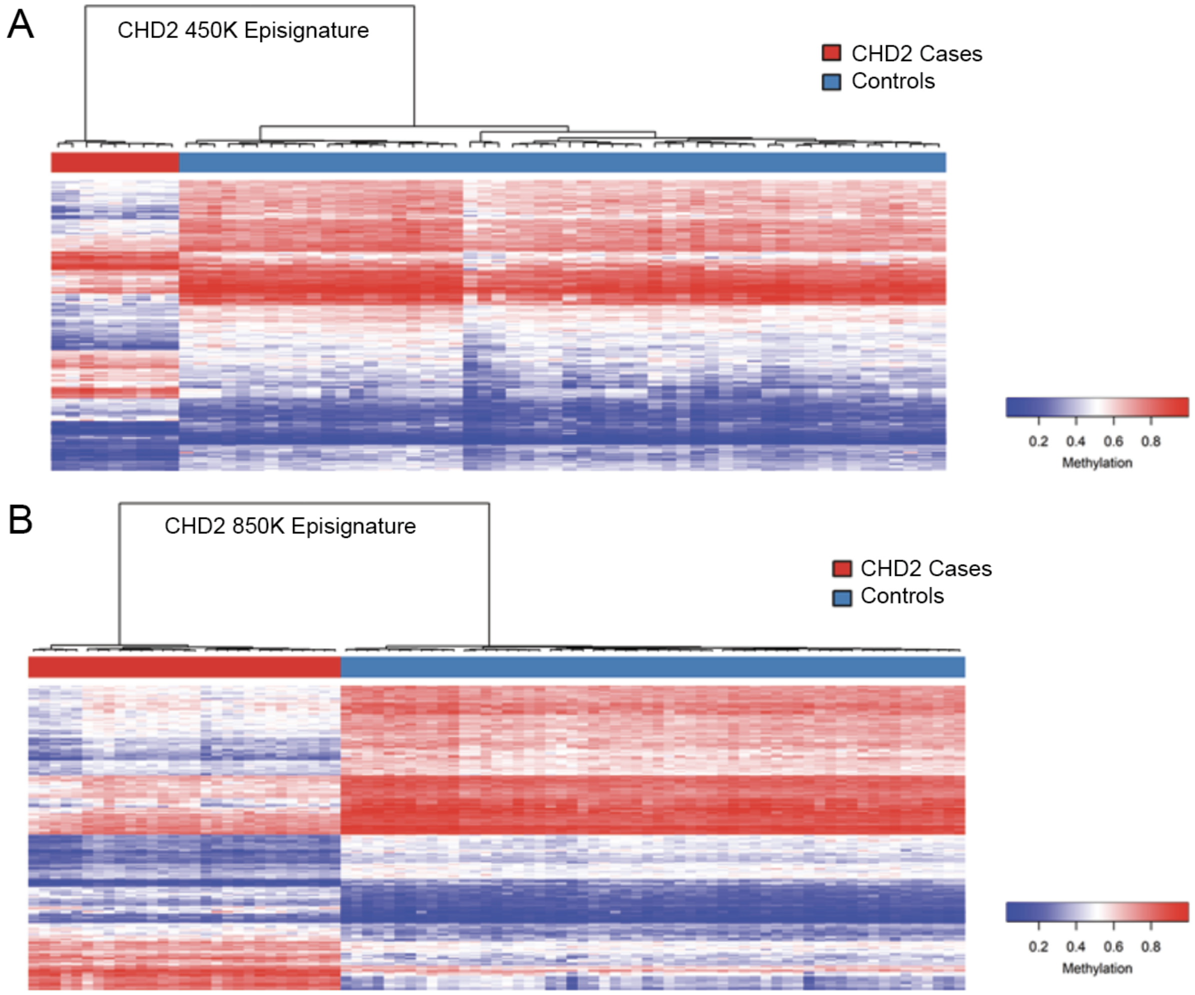


**Figure S21**. Refinement of the *CHD2* Episignature on the 850K array. **A**. Heatmap and dendrogram showing clustering using the *CHD2* 450K episignature probes (n=200), derived using overlapping 450K/850K probes for n=9 individuals (n=2 450K and n=7 850K) with pathogenic variants in *CHD2* against n=54 controls. **B**. Heatmap and dendrogram showing clustering using the *CHD2* 850K episignature probes (n=200), derived using n=29 individuals with pathogenic variants in *CHD2* against n=58 controls.

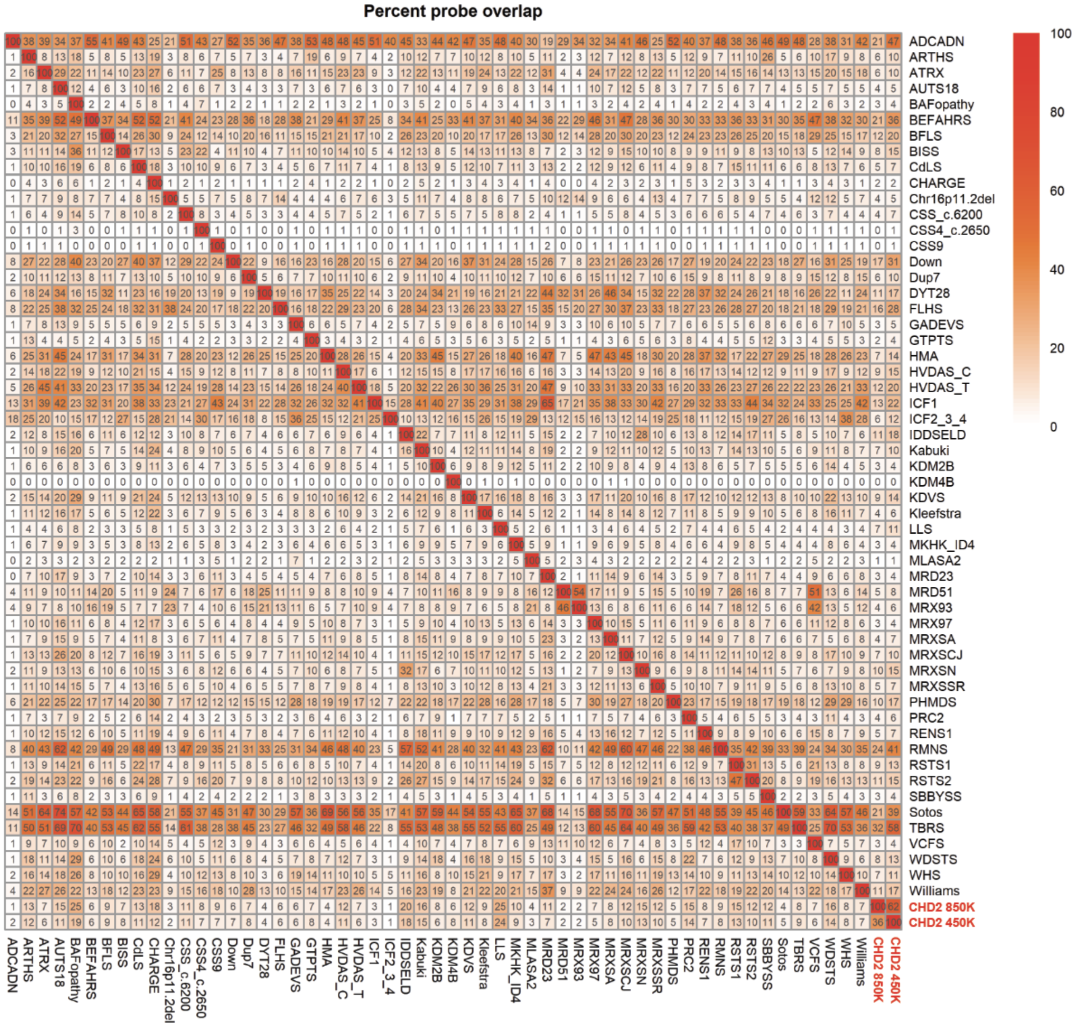


**Figure S22**: Differentially methylated probes found to be shared between multiple cohorts. Percent of probes with an adjusted $p<0.01$ that are shared between each pair of cohorts that was previously investigated[26]. For each pair, the colors indicate the percent of the bottom cohort's probes that are also found in the right cohort's probes. The CHD2 850K episignature shares some portion of probes with every other episignature represented except for the 3 episignatures with the smallest number of DMPs (<500 DMPs): KDM4B/*KDM4B* (MIM:619320), CSS9/*SOX11* (MIM:615866), and CSS4_c.2650/*SMARCA4*, a secondary signature for variants near SMARCA4:c.2650, which cluster separately from other CSS4/BAFopathy (MIM:614609) samples.

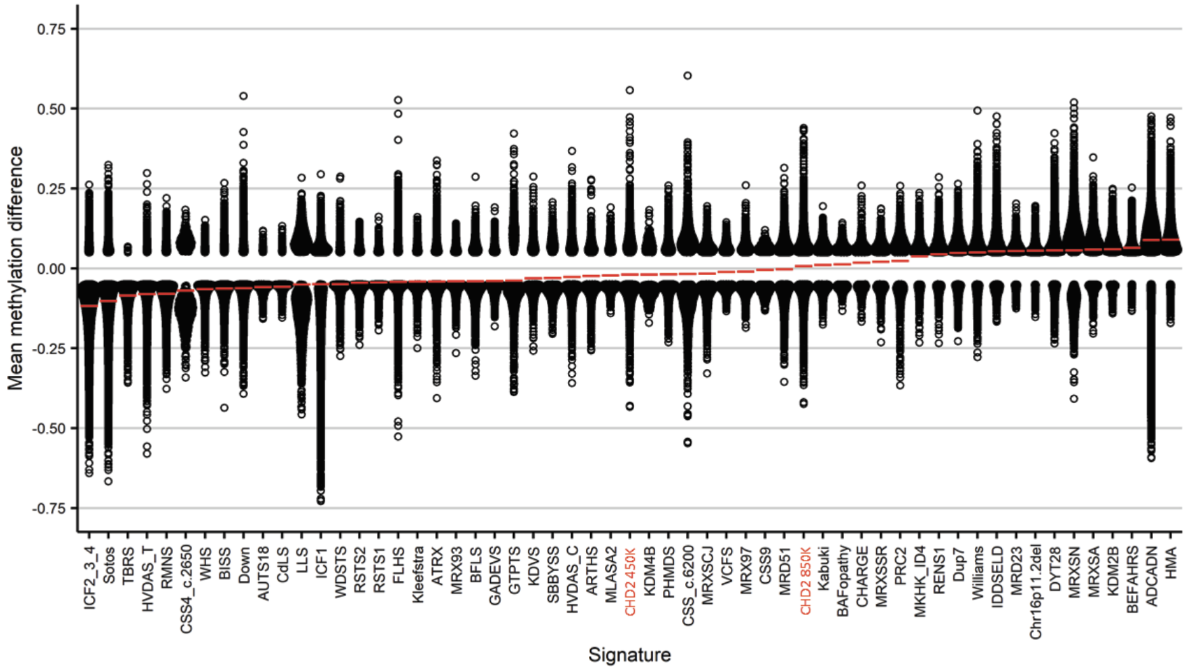


**Figure S23**. Mean Methylation Difference Comparison Across Episignatures. Genome-wide DNA methylation profiles of the updated 850K CHD2 cohort and 55 EpiSign$^{TM}$ episignatures previously investigated for functional correlational analysis[26]. Global methylation differences of all differentially methylated probes (false discovery rate<0.05) for each cohort, sorted by mean methylation. Each circle represents 1 probe. Red lines indicate mean methylation. The x-axis represents 1 of the 57 episignatures and the y-axis is the mean methylation difference.

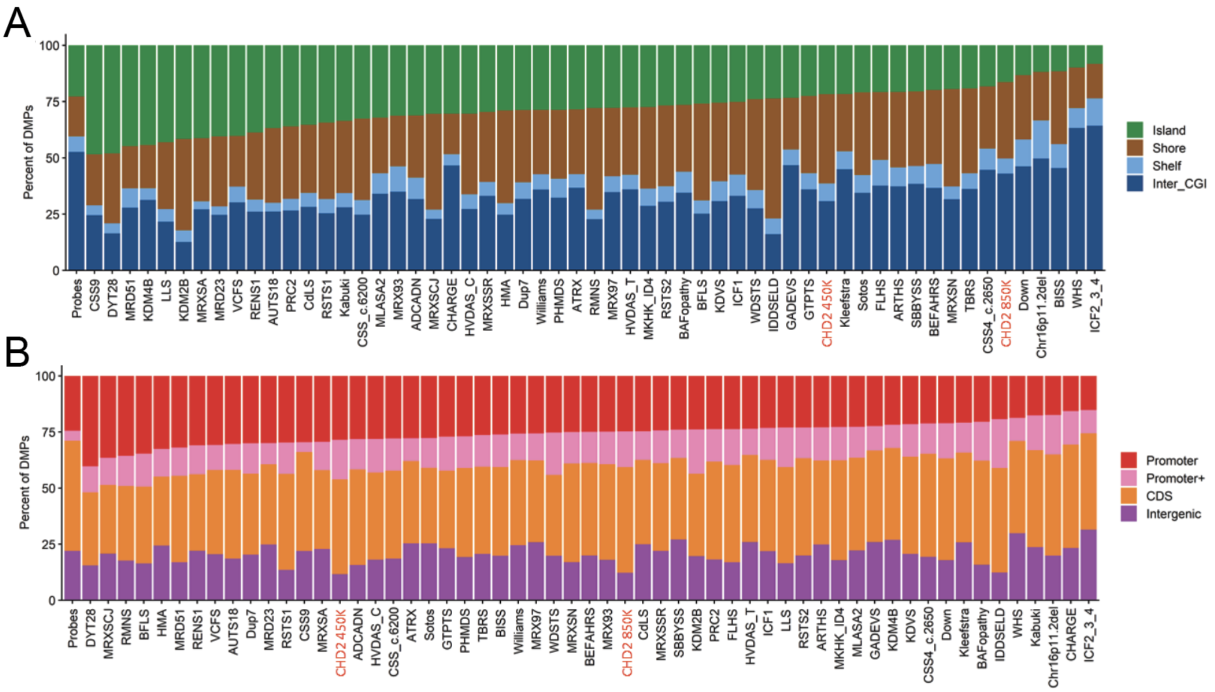


**Figure S24**. Annotation of differentially methylated probes in 450K and 850K CHD2 episignatures and other EpiSign™ episignatures previously published. Differentially methylated probes (DMPs) of CHD2 450K and 850K cohorts (red) and 56 other episignatures as previously investigated[26]. X-axis represents 1 of the 57 episignatures and the y-axis the percentage of DMPs. **A**. DMPs annotated in the context of CpG islands. Island, CpG islands; Shore, within 0 to 2 kb of a CpG island boundary; Shelf, within 2 to 4 kb of a CpG island boundary; inter-CGI, all other regions in the genome **B**. DMPs annotated in the context of genes. Promoter,0 to 1 kb upstream of the TSS; Promoter+, 1 to 5 kb upstream of the TSS; CDS, coding sequence; Intergenic, all other regions of the genome. The Probes column in panels A and B represents the background distribution determined in a previous study[27] of all array probes after initial filtering and used as input for DMP analysis.

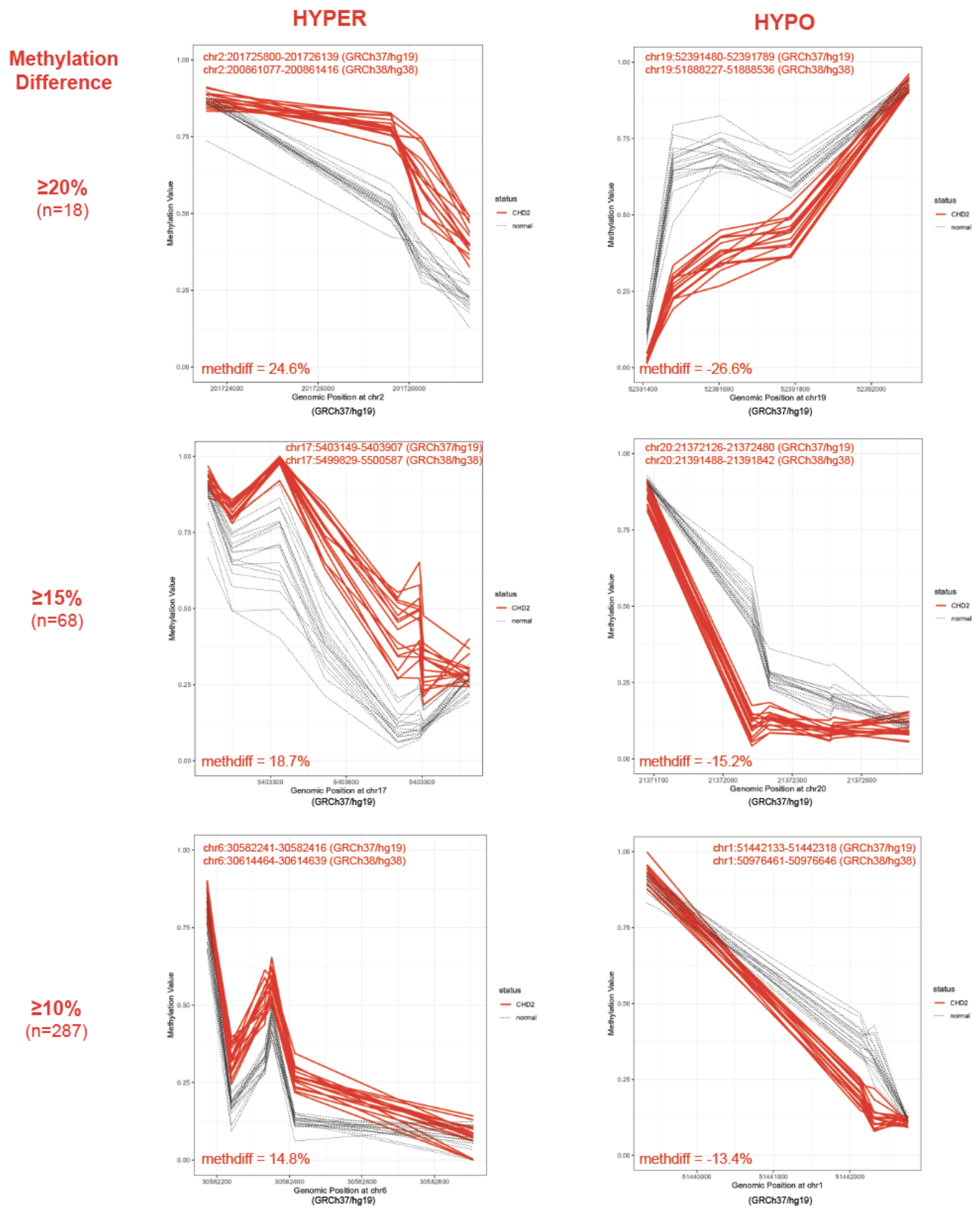


**Figure S25**. Representative *CHD2* DMRs derived on the 850K array. DMRs were called from bumphunter and DMRcate for n=16 individuals with pathogenic variants in *CHD2* versus n=18. Final DMR coordinates were considered the overlapping regions of at least 50bp of more. Representative hypermethylated DMR plots (GRCh37/hg19) are shown on the left, and hypomethylated DMR plots are shown on the right. The methylation differences were considered the average between the two callers and are shown in the DMR plots. The number of DMRs called at various methylation differences are noted to the left of the plots. A total of 712 DMRs were called with at least a 5% methylation difference between *CHD2* and controls.

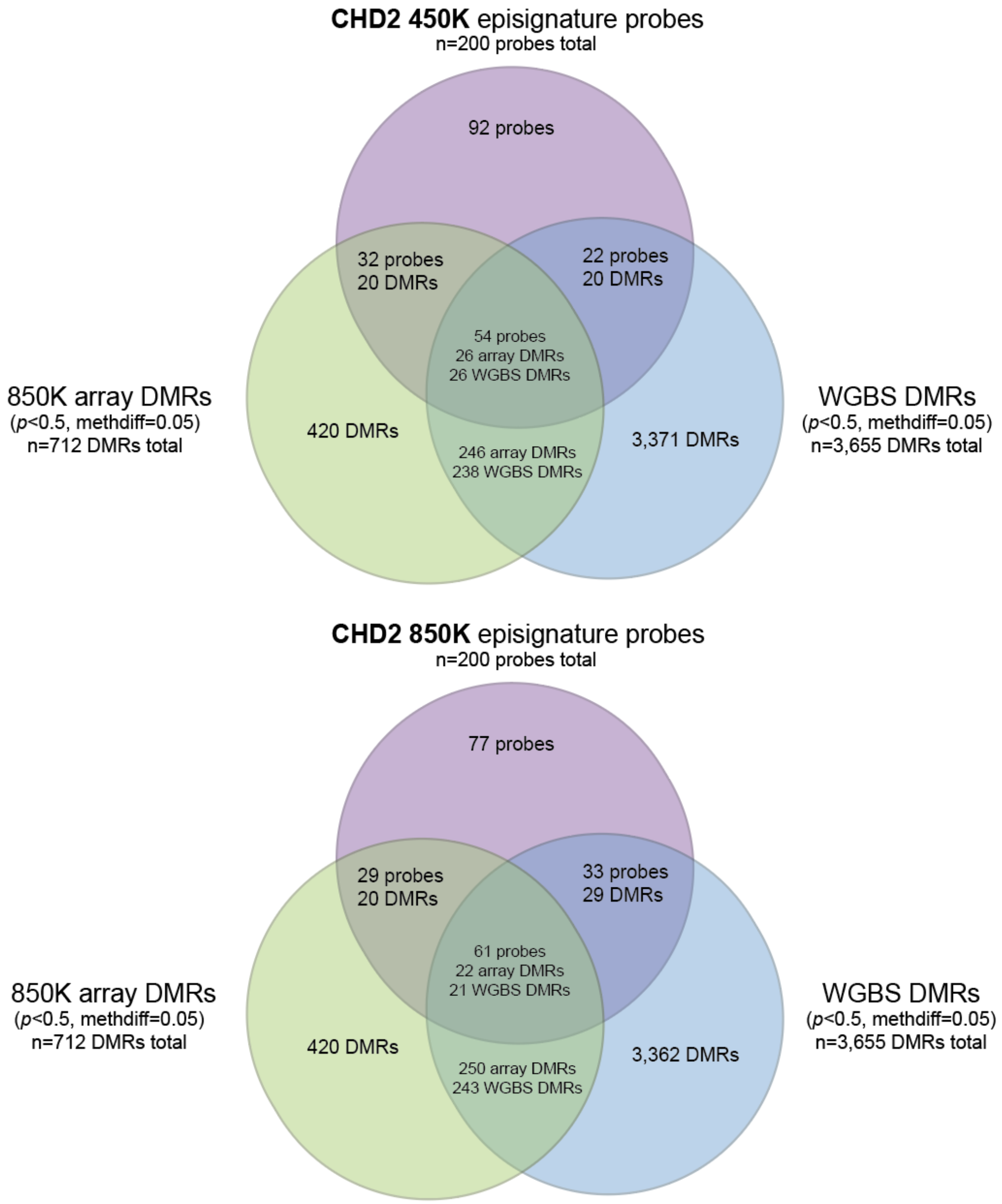


**Figure S26.** Overlap of *CHD2* episignature probes with DMRs called from the 850K array and WGBS. Venn diagrams showing overlap of 450K CHD2 episignature (n=200 probes, top purple) and 850K (n=200 probes, bottom purple) with 850K array DMRs (left green) and WGBS DMRs (right blue). See methods for a detailed description of how episignatures and DMRs were derived.

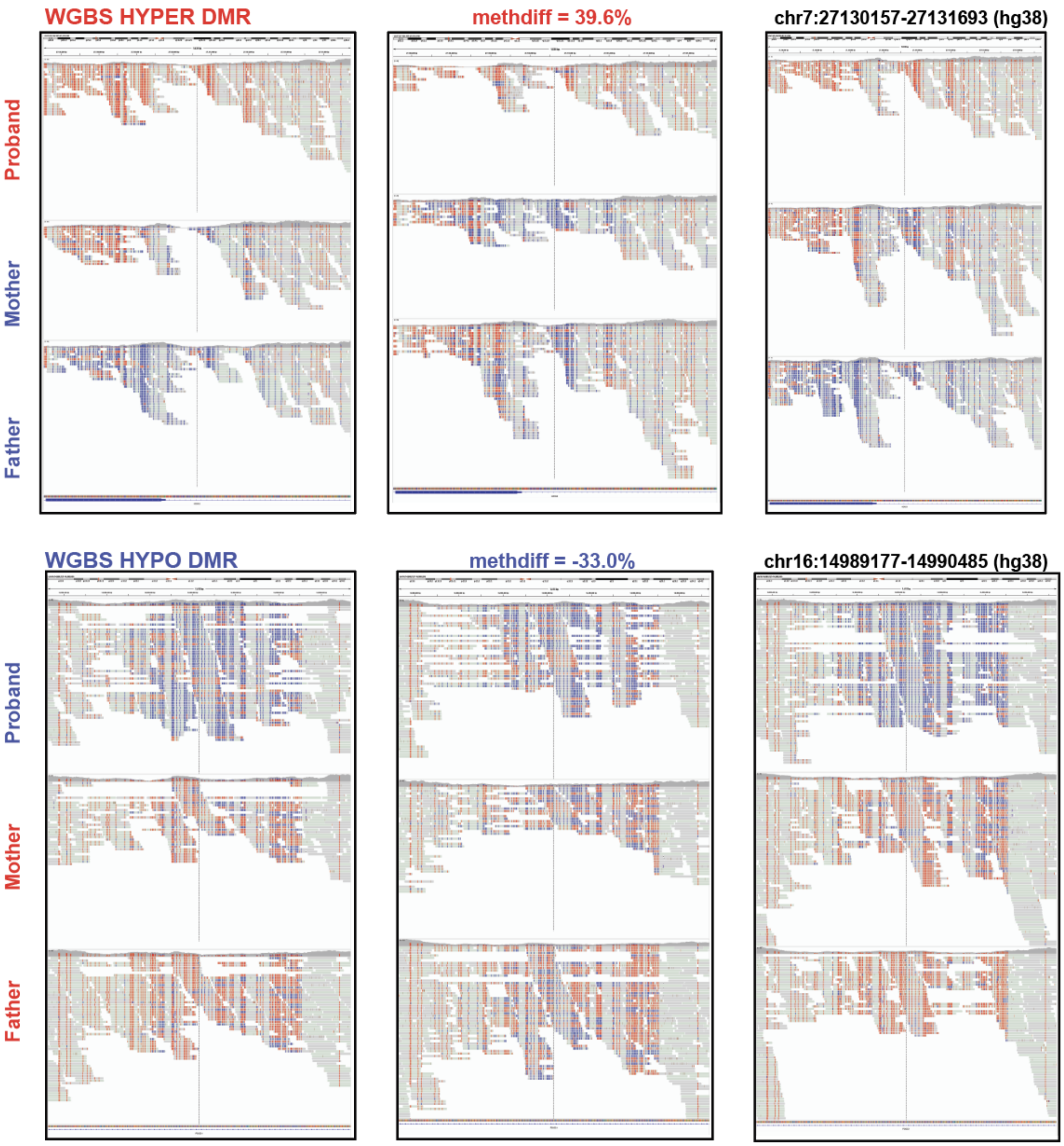


**Figure S27.** Representative *CHD2* DMRs called from WGBS (n=4 probands vs. n=6 parents). DMRs were called for n=16 individuals with pathogenic variants in *CHD2* versus n=18 controls requiring overlap from both DSS and DMRcate tools. Two representative DMRs (1 hyper, 1 hypo) are shown in IGV for n=3 trios. The proband (top track) is clearly differentially methylated compared to both parents (middle and bottom tracks). The center of the plot is marked by a dotted balck line.

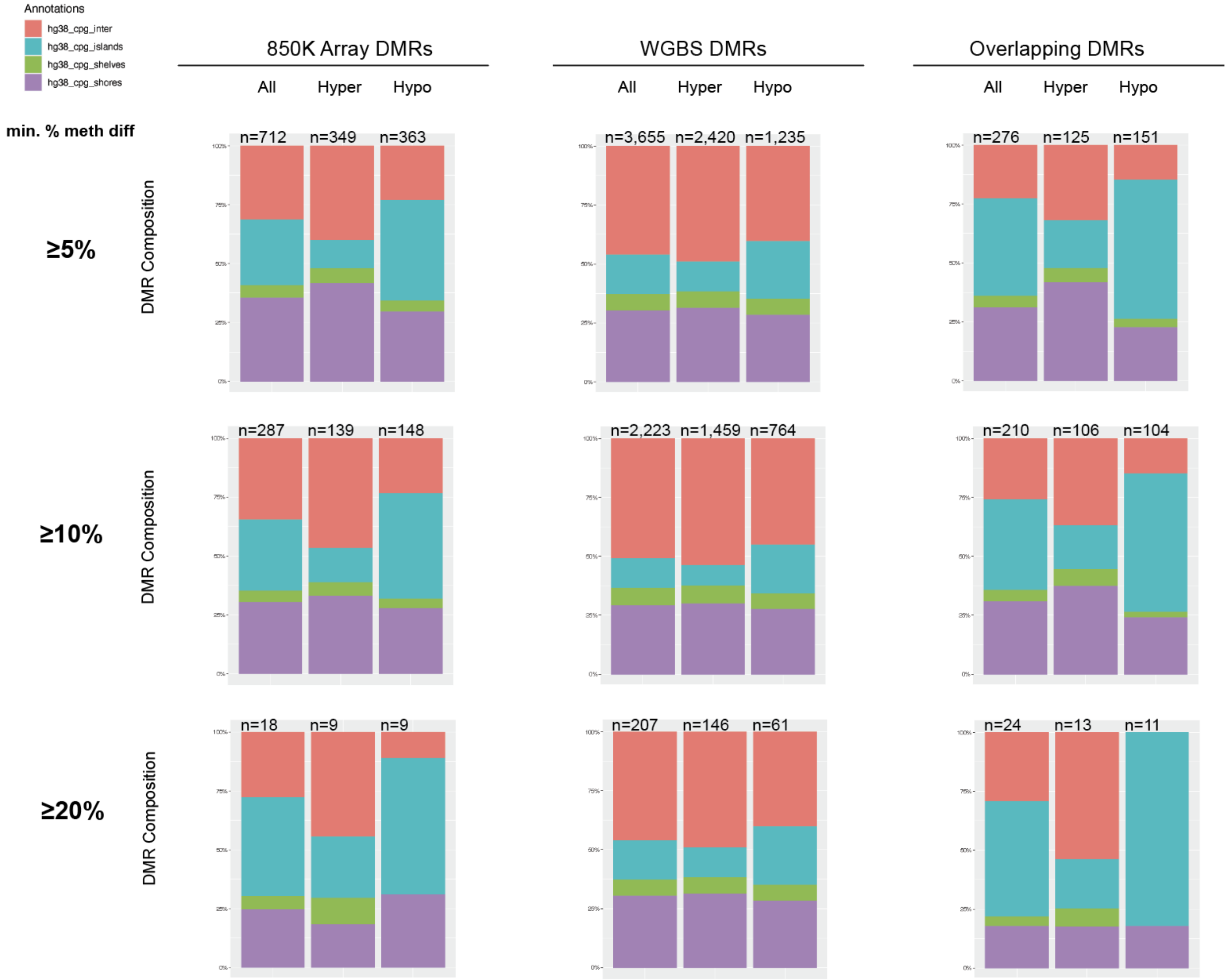


**Figure S28.** Functional annotation of *CHD2* DMRs with CpG descriptions. DMRs were called for n=16 *CHD2* vs. n=18 unaffected controls using bumphunter and DMRcate for the 850K array and DSS and DMRcate for WGBS (Methods). DMRs were considered significant if $p<0.05$ and the percent methylation difference (% meth diff) was greater than or equal to the cutoffs shown (5%, 10%, 20%). DMR composition by CpG descriptors was calculated as a fraction of the length of each DMR to ensure that a DMR overlapping with multiple descriptors was only counted once. CpG islands are areas of the genome with a high density of CpGs. CpG shores are located within 2Kb on either side of CpG islands. CpG shelves are located within 2Kb of CpG shores, and interCpG islands (interCGI) covers the rest of the genome.

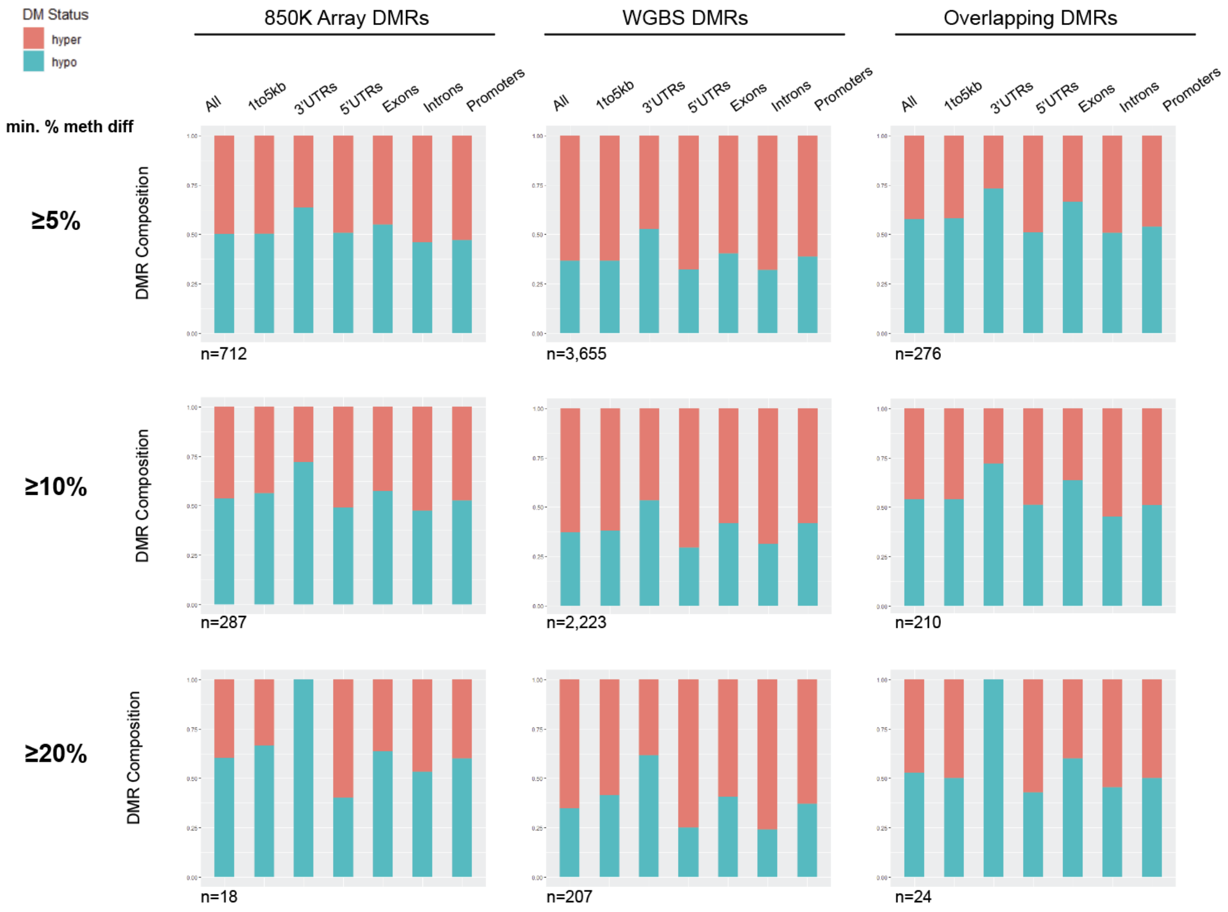


**Figure S29.** Functional annotation of *CHD2* DMRs by gene region. DMRs were called for n=16 *CHD2* vs. n=18 unaffected controls using bumphunter and DMRcate for the 850K array and DSS and DMRcate for WGBS (Methods). DMRs were considered significant if $p<0.05$ and the percent methylation difference (% meth diff) was greater than or equal to the cutoffs shown (5%, 10%, 20%). DMR composition by gene region includes 1-5Kb upstream of the TSS (1to5kb), the promoters (<1Kb upstream the TSS), the 5'UTRs, exons, introns, and the 3'UTRs. Representation as a fraction across hypermethylated DMRs are shown in pink and hypomethylated DMRs are shown in blue.

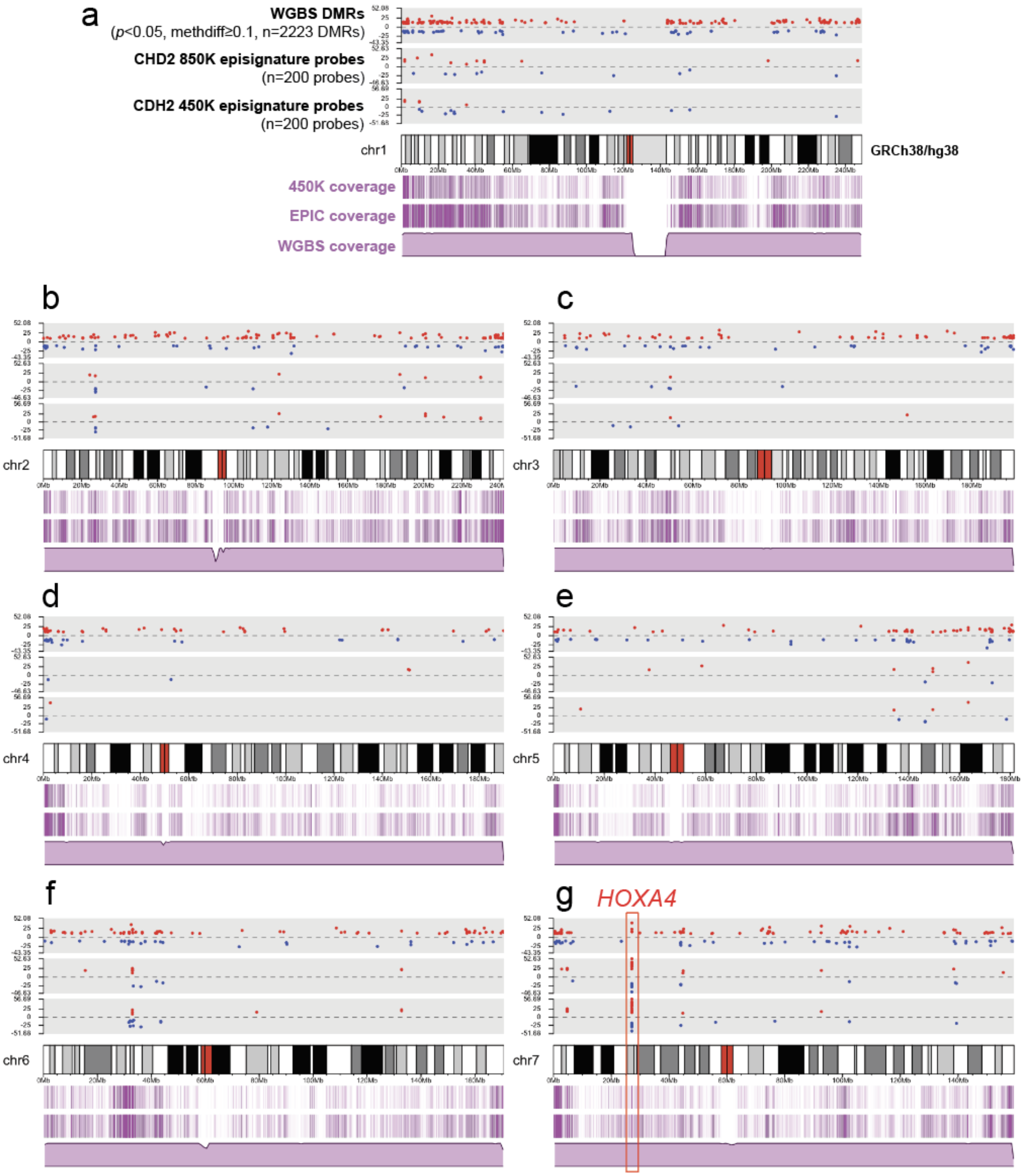

a
WGBS DMRs
(p<0.05, methdiff≥0.1, n=2223 DMRs)
CHD2 850K episignature probes
(n=200 probes)
CDH2 450K episignature probes
(n=200 probes)
chr1
GRCh38/hg38
450K coverage
EPIC coverage
WGBS coverage
b
chr2
c
chr3
d
chr4
e
chr5
f
chr6
g
HOXA4
chr7

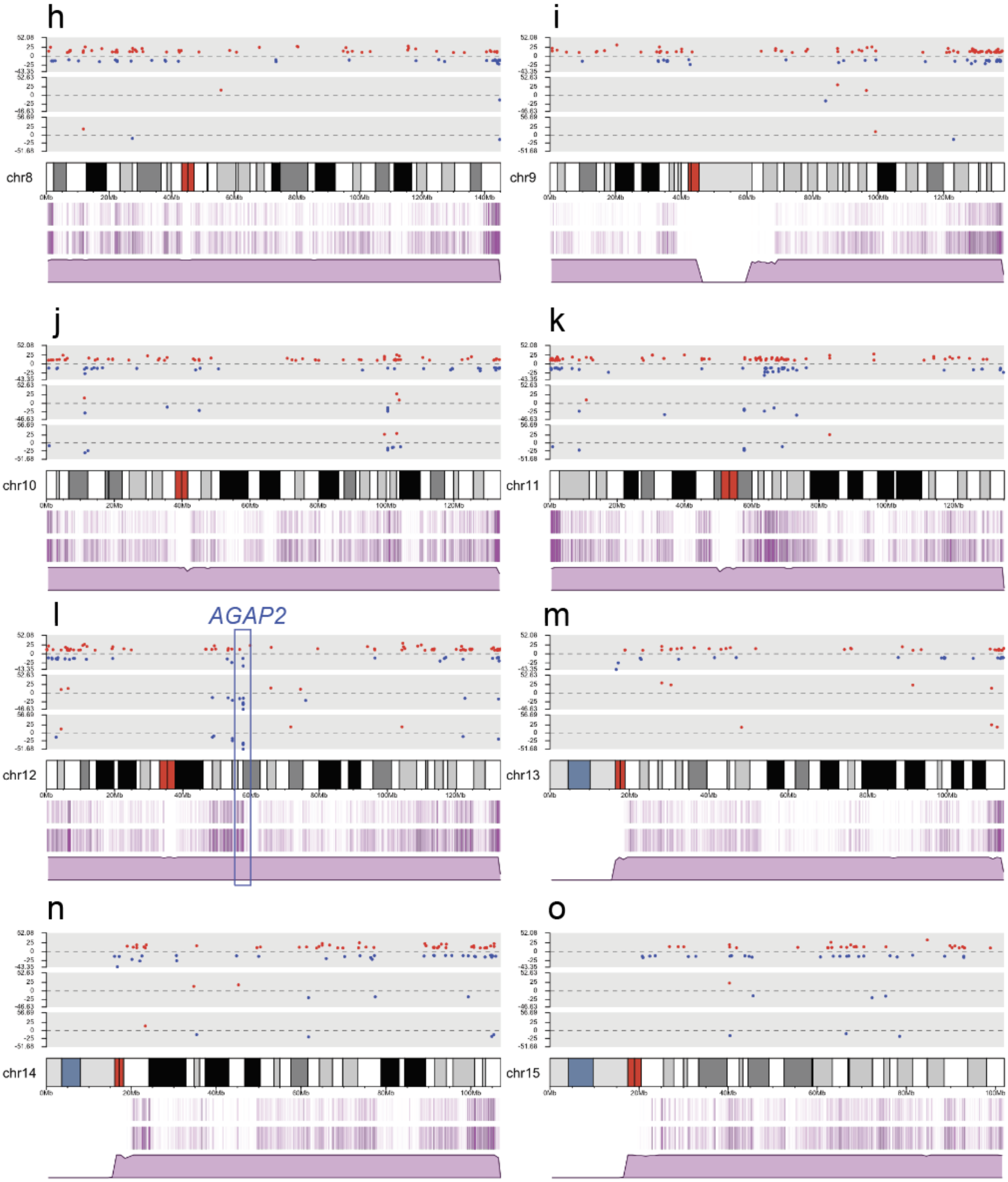
h
chr8
i
chr9
j
chr10
k
chr11
l
AGAP2
chr12
m
chr13
n
chr14
o
chr15

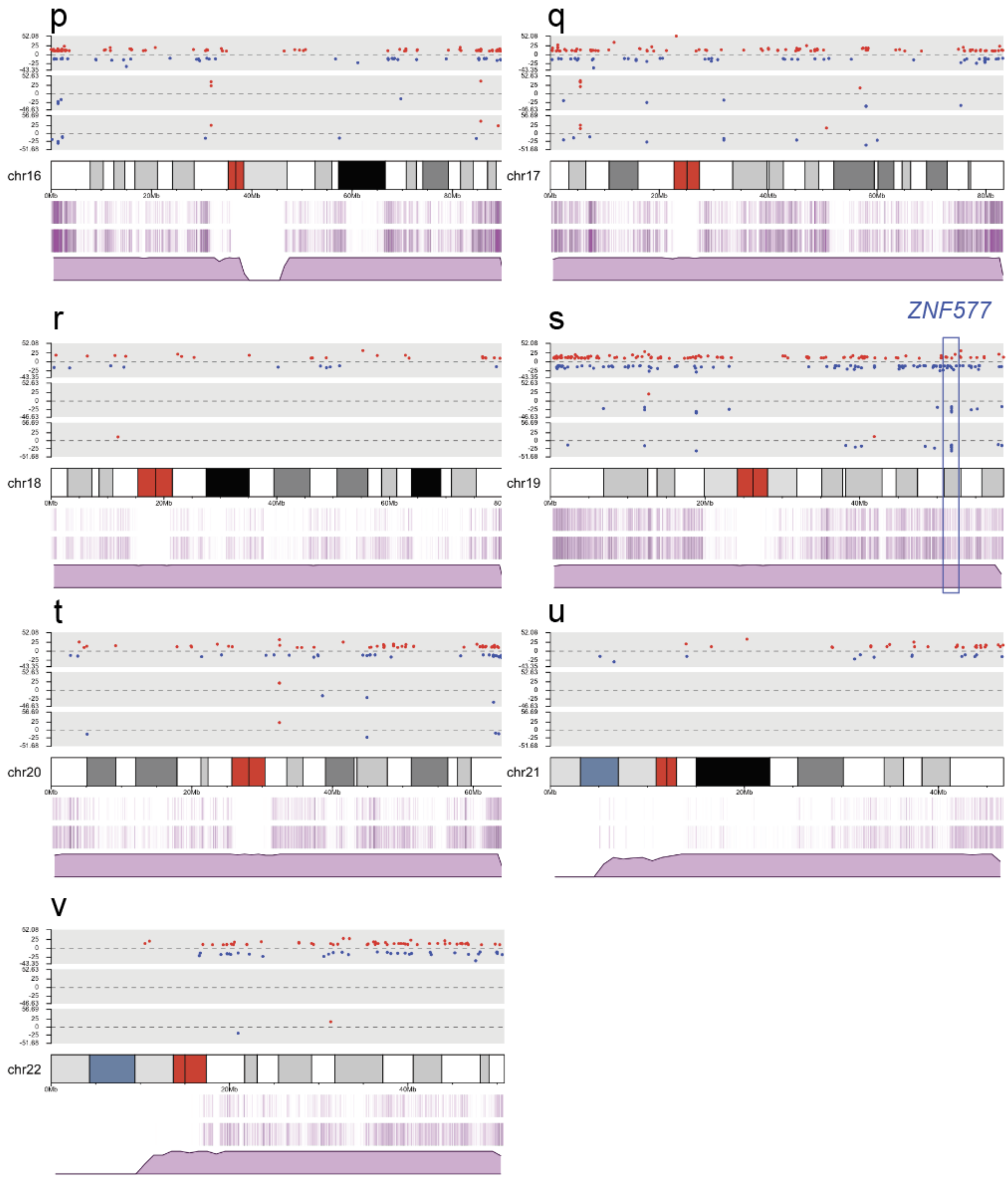


**Figure S30.** Linear karyotype plots representing *CHD2* episignature probes overlapping with DMRs. Individual karyotype tracks for chr1-22 (**a-v**) where three grey tracks (upper panel above chromosome) depict individual red (hyper) or blue (hypo) dots for WGBS DMRs (upper), CHD2

850K episignature probes (middle) and CHD2 450K episignature probes (lower). The scale denotes the methylation difference between *CHD2* relative to controls. Three purple tracks (lower panel below chromosome) depict the coverage for the 450K array probes (lines, upper), 850K array probes (lines, middle), and WGBS reads (distribution, lower). The coverage track for the WGBS is taken from a representative sample after inspecting the average coverage values across all the samples. Examples of where episignature probes form a cluster and overlap with a DMR are boxed in red (hyper) and blue (hypo) for *HOXA4*, *AGAP2*, and *ZNF577*.

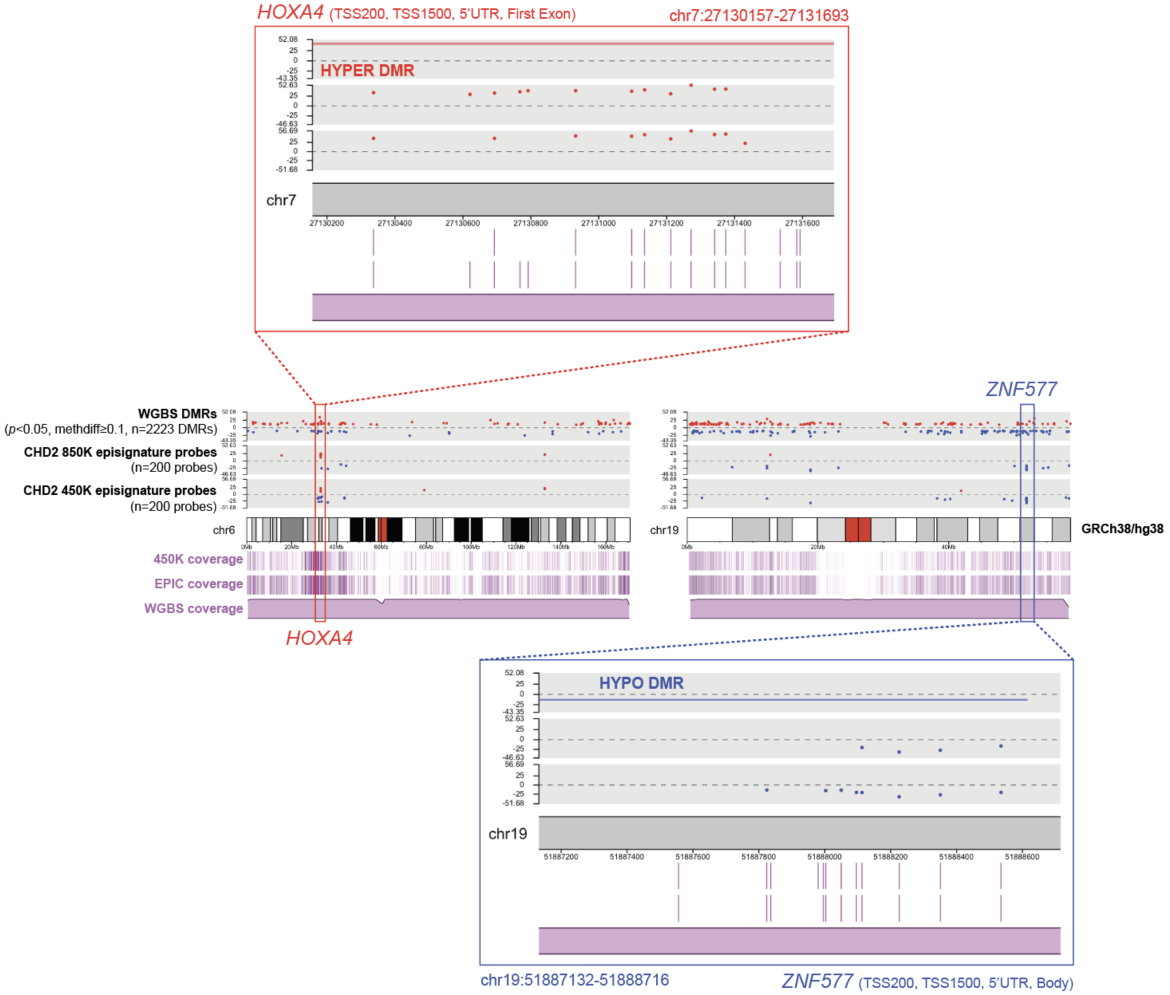


**Figure S31**: Direct overlap of multiple *CHD2* episignature probes with DMRs. "Zoomed" in karyotype plots of regions boxed in Figure S30 for *HOXA4* and *ZNF577*. DMR called from WGBS is depicted as a line in the top grey panel showing total overlap with *CHD2* 450K (bottom grey) and 850K episignature (middle grey) probes. Gene annotations are noted as within 200bp or 1500bp of the transcription start site (TSS200, TSS1500), the 5' untranslated region (5'UTR), or gene body (Body).

### Supplemental Tables Descriptions

**Table S1**: Cohort details, methylation array quality control and filtering numbers.

**Table S2**: Detailed variant table for rare DMRs and episignatures.

**Table S3**: List of all rare outlier DMRs for the autosomes and chrX.

**Table S4**: Comparison of CpG coverage for DMRs validated by targeted EM-seq.

**Table S5**: Variant table from trio sequencing analysis for individual with X;13 translocation.

**Table S6**: X-inactivation assay for parent with *BCLAF3* hypermethylation.

**Table S7**: Probe information for CHD2 450K and 850K episignatures.

**Table S8**: CHD2 DMR master list.

*For access to this data, please contact the corresponding author.*
